# Development of a Simplified Smell Test to Identify Patients with Typical Parkinson’s as Informed by Multiple Cohorts, Machine Learning and External Validation

**DOI:** 10.1101/2024.08.09.24311696

**Authors:** Juan Li, Kelsey Grimes, Joseph Saade, Julianna J. Tomlinson, Tiago A. Mestre, Sebastian Schade, Sandrina Weber, Mohammed Dakna, Tamara Wicke, Elisabeth Lang, Claudia Trenkwalder, Natalina Salmaso, Andrew Frank, Tim Ramsay, Douglas Manuel, aSCENT-PD Investigators, Brit Mollenhauer, Michael G. Schlossmacher

**Affiliations:** Neuroscience Program, Ottawa Hospital Research Institute, Ottawa, ON., Canada; Methodological, Implementation Research Program, Ottawa Hospital Research Institute, Ottawa, ON., Canada; the Methods Centre, Ottawa Hospital Research Institute, Ottawa, ON., Canada; University of Ottawa Brain and Mind Research Institute, Ottawa, ON., Canada; Department of Cellular and Molecular Medicine, University of Ottawa, Ottawa, ON, Canada; Department of Medicine, University of Ottawa, Ottawa, ON, Canada; School of Epidemiology and Public Health, Faculty of Medicine, University of Ottawa, Ottawa, ON, Canada; Division of Neurology, Department of Medicine, The Ottawa Hospital, Ottawa, ON., Canada; Paracelsus-Elena-Klinik, Kassel, Germany; Department of Neurology, University Medical Center Goettingen, Germany; Department of Neuroscience, Carleton University, Ottawa, ON, Canada; Memory Program, Bruyère Research Institute, Ottawa, ON, Canada; Deutsches Zentrum für Neurodegenerative Erkrankungen (DZNE), Goettingen, Germany; Aligning Science Across Parkinson’s (ASAP) Collaborative Research Network, Chevy Chase, MD 20815

**Keywords:** Parkinson disease, parkinsonism, olfaction, hyposmia, smell test, Sniffin’ Stick Test, University of Pennsylvania Smell Identification Test, dementia

## Abstract

**Background:** Reduced olfaction is a common feature of patients with typical Parkinson disease (PD). We sought to develop and validate a simplified smell test as a screening tool to help identify PD patients and explore its differentiation from other forms of parkinsonism.

**Methods:** We used the Sniffin’ Sticks Identification Test (SST-ID) and the University of Pennsylvania Smell Identification Test (UPSIT), together with data from three case-control studies, to compare olfaction in 301 patients with PD or dementia with Lewy bodies (DLB) to 36 subjects with multiple system atrophy (MSA), 32 individuals with progressive supranuclear palsy (PSP) and 281 neurologically healthy controls. Individual SST-ID and UPSIT scents were ranked by area under the receiver operating characteristic curve (AUC) values for group classification, with 10-fold cross-validation. Additional rankings were generated by leveraging results from eight published studies, collectively including 5,853 unique participants. Lead combinations were further validated using (semi-)independent datasets. An abbreviated list of scents was generated based on those shared by SST-ID and UPSIT.

**Findings:** We made the following five observations: (i) PD and DLB patients generally had worse olfaction than healthy controls, as published, with scores for MSA and PSP patients ranking as intermediate. (ii) SST-ID and UPSIT scents showed distinct discriminative performances, with the top odorants (licorice, banana, clove, rose, mint, pineapple and cinnamon) confirmed by external evidence. (iii) A subset of only seven scents demonstrated a similar performance to that of the complete 16-scent SST-ID and 40-scent UPSIT kits, in both discovery and validation steps. Seven scents distinguished PD/DLB subjects from healthy controls with an AUC of 0.87 (95%CI 0.85-0.9) and PD/DLB from PSP/MSA patients with an AUC of 0.73 (95%CI 0.65-0.8) within the three cohorts (n=650). (iv) Increased age was associated with a decline in olfaction. (v) Males generally scored lower than females, although this finding was not significant across all cohorts.

**Interpretation:** Screening of subjects for typical Parkinson’s-associated hyposmia can be carried out with a simplified scent identification test that relies on as few as seven specific odorants. There, the discrimination of PD/DLB subjects vs. age-matched controls is more accurate than that of PD/DLB vs. PSP/MSA patients.

**Funding:** This work was supported by: Parkinson Research Consortium; uOttawa Brain & Mind Research Institute; and the Aligning Science Across Parkinson’s Collaborative Research Network.

**Research in context:** *Evidence before this study:* Chronic hyposmia is a common feature of Parkinson disease (PD) and dementia with Lewy bodies (DLB), which often precedes motor impairment and cognitive dysfunction by several years; it is also frequently associated with α-synuclein aggregate formation in the bulb. The presence of hyposmia increases an individual’s likelihood of having -what has recently been proposed as- a neuronal synucleinopathy disease, by >24-fold. Despite the strong association of PD with reduced olfaction, little is understood about it clinically, such as whether it is affected by sex and age, and whether hyposmia of PD is associated with the same scent identification difficulty seen in other conditions that present with parkinsonism. Moreover, due to its time-consuming nature and traditional administration by healthcare workers, extensive olfactory testing is not routinely performed during neurological assessments in movement disorder clinics.

*Added value of this study:* We analyzed the performance of both the Sniffin’ Sticks Test kit and UPSIT battery to discriminate between healthy controls, patients with PD/DLB and those with MSA or PSP. Comparison to and juxtaposition with eight other published studies allowed for the generation of a markedly abbreviated smell identification test that unified both tests, as described. Group classification performance by each scent and its distractors was further analyzed using machine learning and advanced Item Response Theory methods. Relations between each scent tested, sex and age were analyzed for the first time. Our findings suggest concrete steps to be implemented that would allow for simplified, routine olfaction testing in the future.

*Implications of all the available evidence:* Olfaction testing has emerged as an important neurological assessment part when examining subjects with Parkinson’s and those at risk of it. A simple, validated smell test containing fewer scents than current options could facilitate rapid testing of olfaction in clinic settings and at home, without supervision by healthcare workers. The usefulness of such a non-invasive test in population health screening efforts could be further enhanced when coupled to a self-administered survey that includes questions related to other risk factors associated with PD. As such, large-scale community screening and applications to routine practice in family doctors’ offices as well as in specialty clinics could be made operationally feasible and cost-effective.

## INTRODUCTION

Hyposmia is a common non-motor sign of Parkinson disease (PD) and dementia. The reported prevalence of olfaction loss in PD ranges from 45% to >90% based on populations selected, testing methods, and threshold criteria.^[1]^ Chronic hyposmia is also described as predictive, with reduced olfaction preceding PD diagnosis by 4-20 years.^[1]-[3]^ Olfactory testing may also help in the differentiation of parkinsonian syndromes.^[4]^ Several screening tools and predictive models for the incidence of PD have included subjective or objective assessments of olfaction.^[5]-[9]^

Two commonly used smell tests for evaluating olfactory function include the University of Pennsylvania Smell Identification Test (UPSIT),^[10]^ a battery of 40 scratch-and-sniff questions that are self-administered, and the Sniffin’ Sticks Test (SST) battery,^[11],[12]^ with three subtests (for Identification; Discrimination; Threshold) comprising 16 scents each, of which the administration usually involves a trained supervisor. The SST-Identification (SST-ID) and UPSIT are comparable in that they both assess one’s ability to identify a range of scents. Within SST, SST-ID is more commonly used in PD cohort studies and known to have better diagnostic performance than the SST-Discrimination subtest.^[13]^

Smell test kits were initially developed to assess olfaction in the general population but have been increasingly used in research settings that study disorders of the brain. Using different cohorts and methods, some studies have ranked odorants in the UPSIT^[14]-[17]^ and SST-ID kits^[13],[18]-[20]^ by their diagnostic performances, and reported that certain subsets of scents appeared to have equal or better performance than the entire complement of 40- or 16-scents-based tests. Other studies^[21]-[24]^ have examined subsets of UPSIT but without any details on scent ranking. Moreover, external validation was frequently missing in these analyses, proposed scent combinations were found to be cohort-specific without agreement across different studies,^[16],[25]^ and the role of distractors (versus the correct scent offered) in such multi-choice settings were understudied. Furthermore, analyses of UPSIT and SST-ID kits were always conducted separately despite the similarities between the two tests. Finally, olfaction scores in patients with other, atypical forms of parkinsonism were not assessed in PD-centric studies.

In the current work, we aimed to assess olfaction performances in commonly encountered forms of parkinsonism, to assess the key features of both UPSIT and SST-ID odorants, to explain any observed differences between them, and to develop a simplified smell test to unify both kits using proper internal and external validation steps. To this end, eight published scent rankings,^[13]-[20]^ which collectively examined 5,853 participants, were incorporated into our study to make the proposed abbreviated test generalizable and to avoid overfitting. We also added Item Response Theory-based analyses when examining the behaviour of participants’ responses to multiple choices provided for each scent; this, to enhance sensitivity and specificity of future test versions. Lastly, we analyzed the effects of age and sex on performance in scent identification.

## METHODS

The study was conducted in adherence with STARD^[26]^ guideline, see **Supplemental Table 1**.

### Source of data and participants

We used de-identified data from three observational, retrospective, case-control studies: the “*De Novo* Parkinson disease study” (DeNoPa);^[27]^ Ottawa (PREDIGT) Trial; and Prognostic Biomarkers in Parkinson’s Disease Study (PROBE).^[28]^ Detailed description and inclusion/exclusion criteria of these three cohorts are provided in **Supplemental Methods**; their demographic and diagnostic characteristics are summarized in **Table 1**. Data of the cross-sectional Ottawa Trial study, baseline data of the longitudinal PROBE study, and three visits of the longitudinal DeNoPa study (baseline; 48-month; and 72-month follow-up visits) were used. Patients with PD or dementia with Lewy bodies (DLB) (DeNoPa: n=129; Ottawa Trial: n=70; PROBE: n=102), subjects with multiple system atrophy (MSA) or progressive supranuclear palsy (PSP) (DeNoPa: n= 9; Ottawa Trial: n=6; PROBE: n=53), and neurologically healthy controls (HC) (DeNoPa: n=109; Ottawa Trial: n=118; PROBE: n=54) were included. Diagnostic criteria were applied according to international guidelines; ^[27]-[30]^ diagnoses were revised, as necessary, during longitudinal follow up visits or by independent raters who were blind to olfaction test results. Most study participants with PD in the three trials were classified as Hoehn and Yahr stage II-III. No participant overlap existed between the three studies.

**Table 1:** Baseline demographic characteristics and smell test scores of adults enrolled in the three cohorts. DeNoPa = *De Novo* Parkinson Study. PROBE = Prognostic Biomarkers in Parkinson Disease. IQR = interquartile range. HC = healthy control. PD = Parkinson disease. DLB = dementia with Lewy bodies. MSA = multiple system atrophy. PSP = progressive supranuclear palsy. NA = not applicable. ND = not documented

| Variable | DeNoPa |  |  |  |  | Ottawa Trial |  |  |  |  | PROBE |  |  |  |  |
| --- | --- | --- | --- | --- | --- | --- | --- | --- | --- | --- | --- | --- | --- | --- | --- |
|  | HC,<br>N = 109<sup>1</sup> | PD/DLB,<br>N = 129<sup>1,4</sup> | MSA/PSP,<br>N = 9<sup>1,4</sup> | p-value<sup>2</sup> | q-<br>value<sup>3</sup> | HC,<br>N = 118<sup>1</sup> | PD/DLB,<br>N = 70<sup>1,5</sup> | MSA/PSP,<br>N = 6<sup>1,5</sup> | p-value<sup>2</sup> | q-<br>value<sup>3</sup> | HC,<br>N = 54<sup>1</sup> | PD/DLB,<br>N = 102<sup>1,6</sup> | MSA/PSP,<br>N = 53<sup>1,6</sup> | p-value<sup>2</sup> | q-<br>value<sup>3</sup> |
| Sex |  |  |  | 0.6 | 0.7 |  |  |  | 0.005 | 0.015 |  |  |  | 0.058 | 0.087 |
| Female | 42 (39%) | 45 (35%) | 2 (22%) |  |  | 74 (63%) | 29 (41%) | 5 (83%) |  |  | 28 (52%) | 34 (33%) | 18 (34%) |  |  |
| Male | 67 (61%) | 84 (65%) | 7 (78%) |  |  | 44 (37%) | 41 (59%) | 1 (17%) |  |  | 26 (48%) | 68 (67%) | 35 (66%) |  |  |
| Age | 65 (60, 70) | 66 (58, 72) | 72 (65, 76) | 0.073 | 0.15 | 68 (58, 73) | 68 (60, 74) | 66 (63, 70) | 0.9 | 0.9 | 59 (55, 69) | 61 (55, 69) | 67 (61, 75) | 0.002 | 0.007 |
| Parkinsonism duration<br>at baseline in months | NA | 14 (9, 24) | 12 (6, 33) | 0.7 | 0.7 | NA | 84 (36, 132) | 48 (30, 66) | 0.078 | 0.12 | NA | 65 (58, 71) | 58 (57, 60) | 0.2 | 0.2 |
| Follow-up time in<br>months | 120 (120, 120) | 120 (72, 120) | 72 (48, 120) | 0.001 | 0.004 | ND | ND | ND |  |  | ND | ND | ND |  |  |
| Smell test score<sup>7</sup> | 12 (11, 14) | 7 (4, 9) | 10 (9, 11) | <0.001 | <0.001 | 32 (29, 35) | 17 (13, 22) | 29 (26, 31) | <0.001 | <0.001 | 35 (33, 37) | 21 (14, 26) | 28 (21, 32) | <0.001 | <0.001 |
| Smell test percentile<sup>8</sup> | 50 (25, 75) | 4 (4, 10) | 25 (18, 50) | <0.001 | <0.001 | 39 (18, 66) | 6 (4, 10) | 23 (14, 37) | <0.001 | <0.001 | 56 (27, 73) | 5 (4, 17) | 23 (9, 42) | <0.001 | <0.001 |
| Olfaction<sup>9</sup> |  |  |  | <0.001 | <0.001 |  |  |  | <0.001 | <0.001 |  |  |  | <0.001 | <0.001 |
| Normal | 94 (86%) | 30 (23%) | 7 (78%) |  |  | 93 (79%) | 7 (10%) | 3 (50%) |  |  | 50 (93%) | 29 (28%) | 34 (64%) |  |  |
| Hyposmia/Anosmia | 15 (14%) | 99 (77%) | 2 (22%) |  |  | 25 (21%) | 63 (90%) | 3 (50%) |  |  | 4 (7%) | 73 (72%) | 19 (36%) |  |  |

<sup>1</sup> n (%); Median (IQR); <sup>2</sup> Fisher's exact test; Kruskal-Wallis rank sum test; <sup>3</sup> False discovery rate correction for multiple testing <sup>4</sup> PD: n = 126; DLB: n = 3; MSA: n = 4; PSP: n = 5. <sup>5</sup> PD: n = 69; DLB: n = 1; MSA: n = 5; PSP: n = 1. <sup>6</sup> PD: n = 102; MSA: n = 27; PSP: n = 26. <sup>7</sup> SST-ID scores (0-16) for DeNoPa, UPSIT scores (0-40) for Ottawa Trial and PROBE. <sup>8</sup> Age- and sex-adjusted normalized percentiles. <sup>9</sup> Hyposmia/anosmia was determined by SST-ID percentile ≤ 10%, and UPSIT percentile ≤ 15%.

Analyses of these deidentified cohort data were approved by Investigational Review Boards at Paracelsus-Elena-Klinik (Kassel, Germany) in Frankfurt, Hesse (FF 89/2008), at The Ottawa Hospital (Ottawa, Canada; 20180010-01H) as well as all PROBE Study-affiliated sites, with the participants’ consent.

### Study assessments and statistical analyses

#### Sniffin’ Sticks test (SST)

Used in DeNoPa, the SST comprises a supervised test for one’s sense of smell using pen-like odor dispensing devices, as administered in a clinic setting.^[11],[12]^ The test has three subtests: Identification (SST-ID), Discrimination (SST-DS), and Threshold (SST-TH), each with 16 odorants. In SST-ID, subjects are presented a stick and choose the scent from four options (one correct, three distractors). SST-DS is performed using triplets of odorants that are of similar intensity and hedonic tone, where subjects are required to identify which stick of the triplet has a different scent from the other two. SST-TH is performed using triplets of sticks where only one is filled with odorant at a certain dilution whereas the other two are filled with odor-free solvent. SST-TH determines at what dilution subjects can consistently identify the odorant-filled stick. The entire SST (in German) was completed by all DeNoPa participants at their baseline visit, and the SST-ID subtest was re-administered at 48-month and 72-month follow-up visits.

#### The University of Pennsylvania Smell Identification Test (UPSIT)

UPSIT was used in the Ottawa Trial and PROBE; this self-administered kit contains 40 scratch-and-sniff questions; each question has one correct answer and three incorrect distractors.^[10]^

#### Data preparation

Observations that had no valid response to SST-ID or UPSIT were removed from the analysis. Observations with incomplete responses were imputed with 0s, indicating incorrect responses. A dichotomous response-based transformation (0 = incorrect response; 1 = correct response) was used to calculate the sum scores and assess discriminative performances for each scent. The exact indices of chosen options were used for Item Response Theory analysis (see below).

#### Demographic and diagnostic characteristics

Demographic and diagnostic characteristics of the study cohort were summarised using n (%) or median (the interquartile range (IQR)). The reported p-values represented the significance from corresponding Fisher’s exact test or Kruskal-Wallis rank sum test, with false discovery rate correction for multiple testing; p-values smaller than 0.05 were considered significant.

#### Comparing different smell tests

Score distributions of corresponding smell tests in each subject group were illustrated using Cummings estimation plots.^[31]^ The raw UPSIT and SST-ID scores were also normalized into percentiles based on age and sex, and hyposmia was defined by SST-ID percentile ≤ 10%^[32]^ or UPSIT percentile ≤ 15%.^[10]^ Discrimination performances of these subtests were compared using area under the receiver operating characteristic (ROC) curve (AUC) values with bootstrap estimated 95% confidence interval (CI),^[33]^ in order to distinguish diagnostic groups. **Table 2** also reports optimal thresholds and their associated sensitivity and specificity that correspond to the maximum Youden indices.^[34]^

**Table 2:** Discriminative performances of complete smell tests for baseline visits in three cohorts. DeNoPa = *De Novo* Parkinson Study. PROBE = Prognostic Biomarkers in Parkinson Disease. HC = healthy control. PD = Parkinson disease. DLB = dementia with Lewy bodies. MSA = multiple system atrophy. PSP = progressive supranuclear palsy. AUC = area under the ROC curve. CI = confidence interval. SST-ID = Sniffin’ Sticks Identification test. SST-TH = Sniffin’ Sticks Threshold test. SST-DS = Sniffin’ Sticks Discrimination test. UPSIT = University of Pennsylvania Smell Identification Test.

| Cohort | Test | AUC (95% CI) | Threshold | Sensitivity | Specificity |
| --- | --- | --- | --- | --- | --- |
| **PD/DLB vs HC** |  |  |  |  |  |
| DeNoPa | SST-ID | 0.89 (0.85-0.93) | $\leq 10.5$ | 0.88 | 0.81 |
| | SST-TH | 0.83 (0.78-0.88) | $\leq 3.12$ | 0.68 | 0.9 |
| | SST-DS | 0.83 (0.77-0.88) | $\leq 10.5$ | 0.75 | 0.78 |
| Ottawa Trial | UPSIT | 0.92 (0.88-0.96) | $\leq 28.5$ | 0.96 | 0.77 |
| PROBE | UPSIT | 0.93 (0.89-0.97) | $\leq 29.5$ | 0.84 | 0.89 |
| **PD/DLB vs MSA/PSP** |  |  |  |  |  |
| DeNoPa | SST-ID | 0.8 (0.69-0.91) | $\leq 6.5$ | 0.48 | 1 |
| | SST-TH | 0.64 (0.48-0.8) | $\leq 1.38$ | 0.43 | 0.89 |
| | SST-DS | 0.56 (0.32-0.79) | $\leq 12.5$ | 0.88 | 0.33 |
| Ottawa Trial | UPSIT | 0.92 (0.85-0.99) | $\leq 25.5$ | 0.81 | 1 |
| PROBE | UPSIT | 0.69 (0.6-0.78) | $\leq 26.5$ | 0.76 | 0.58 |

#### Machine learning workflow of developing and validating the abbreviated smell test

**Figure 1** illustrates the machine learning workflow of ranking UPSIT/SST-ID scents, developing and validating respective simplified tests and the unified abbreviated test. Data of the Ottawa Trial and baseline data of DeNoPa were used as discovery cohorts, and baseline data of PROBE and follow-up data of DeNoPa were used for (semi-)independent validation.

1. *Ranking the individual scents*: Using the corresponding discovery cohorts with 10-fold cross validation (see **Supplemental Methods**), individual scents in SST-ID and UPSIT were ranked separately based on their AUC values in differentiating PD/DLB patients from healthy controls. To control over-fitting, the SST-ID and UPSIT scent rankings from our study was compared with eight external rankings,^[13]-[20]^ four for each test, and two final lists were generated by averaging internal and external rankings. Eleven scents are shared by both smell tests, and an additional “Shared” ranking was constructed using their respective positions in the averaged SST-ID and UPSIT rankings.
2. *Developing and validating the best-performing, simplified tests*: For each scent ranking, beginning with the highest-ranked odorant, subsets were constructed by adding one scent at a time in descending ranking order. A total of 95 and 220 distinct SST-ID and UPSIT subsets with various numbers of scents were compared, using their AUC values in distinguishing PD/DLB from HC, to develop the best-performing simplified tests, including one that unified both smell tests. The resulted abbreviated smell tests were also validated using (semi-)independent datasets.

**Figure 1:**
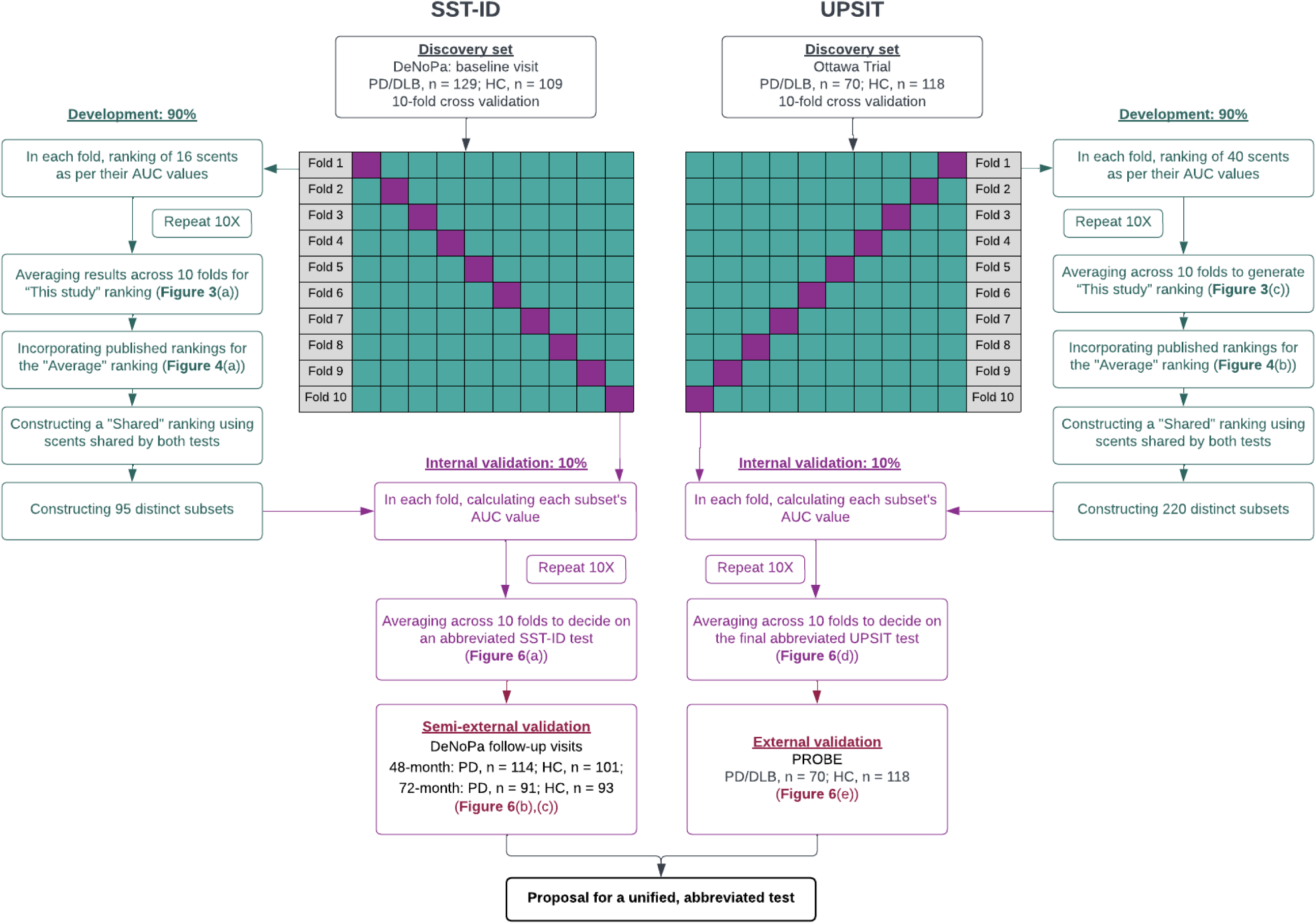
Machine learning workflow for developing and validating an abbreviated smell test. Details of the workflow are as indicated and described in Methods and Result sections of the main text. SST-ID = Sniffin’ Sticks Identification test. UPSIT = University of Pennsylvania Smell Identification Test. DeNoPa = De Novo Parkinson Study. PROBE = Prognostic Biomarkers in Parkinson Disease. HC = healthy control. PD = Parkinson disease. DLB = dementia with Lewy bodies. MSA = multiple system atrophy. PSP = progressive supranuclear palsy. ROC = receiver operating characteristic. AUC = area under the ROC curve.

#### Exploring observed differences in scent performance

The percentage of correct scent identification within each subject group was calculated. These percentages were further compared to examine the relationship of scent identification versus sex and age (using spline smoothing, see **Supplemental Methods**). Furthermore, Item Characteristic Curves (ICCs) for each scent within PD/DLB and HC groups from baseline DeNoPa and Ottawa Trial visits were used to analyze scent performance and the influence of distractors (see **Supplemental Methods** for more details).

Statistical analyses were performed using ‘R’ (version 4.3.1). Cummings estimation plots were generated using ‘dabestr’,^[36]^ and ICC curves were generated using ‘TestGardener’;^[37]^ all other plots were generated using ‘ggplot2’.^[38]^ The package ’pROC’^[33]^ was used for ROC and AUC, and ‘fda’ was used for spline smoothing.^[39]^

## RESULTS

### Comparison of different smell tests for classifying typical Parkinson disease

We first assessed the performance of UPSIT and SST-ID kits in each diagnostic group. As expected, across all three cohorts, PD and DLB patients generally had lower scores (*i.e.*, worse olfaction) than healthy controls (HC), whereas scores of MSA and PSP patients were intermediate; these are shown by score distributions in **Figure 2(a)-(d)**, and median scores/percentiles and percentage of hyposmia in **Table 1**. There was no significant difference in olfaction performance between MSA and PSP patients (**Figure 2(b), (d)**). UPSIT and SST-ID kits had comparable performance in distinguishing PD/DLB patients from HC subjects (**Figure 2(e)**, left) with AUC values in the three cohorts ranging between 0.89-0.93. Further, both tests showed reduced performance when distinguishing PD/DLB patients from MSA/PSP patients (**Figure 2(e)**, right), but with a larger variation (0.69 to 0.92) across the three cohorts, likely due to the small sample sizes of MSA/PSP groups in the Ottawa Trial and DeNoPa cohorts. When compared to the other two SST subtests, SST-ID was found to be the most discriminative one in distinguishing PD/DLB from both HC and from MSA/PSP (**Supplemental Figure 1**), as expected.^[13]^ For cohort-specific thresholds and their corresponding sensitivity and specificity, see **Table 2**.

**Figure 2:**
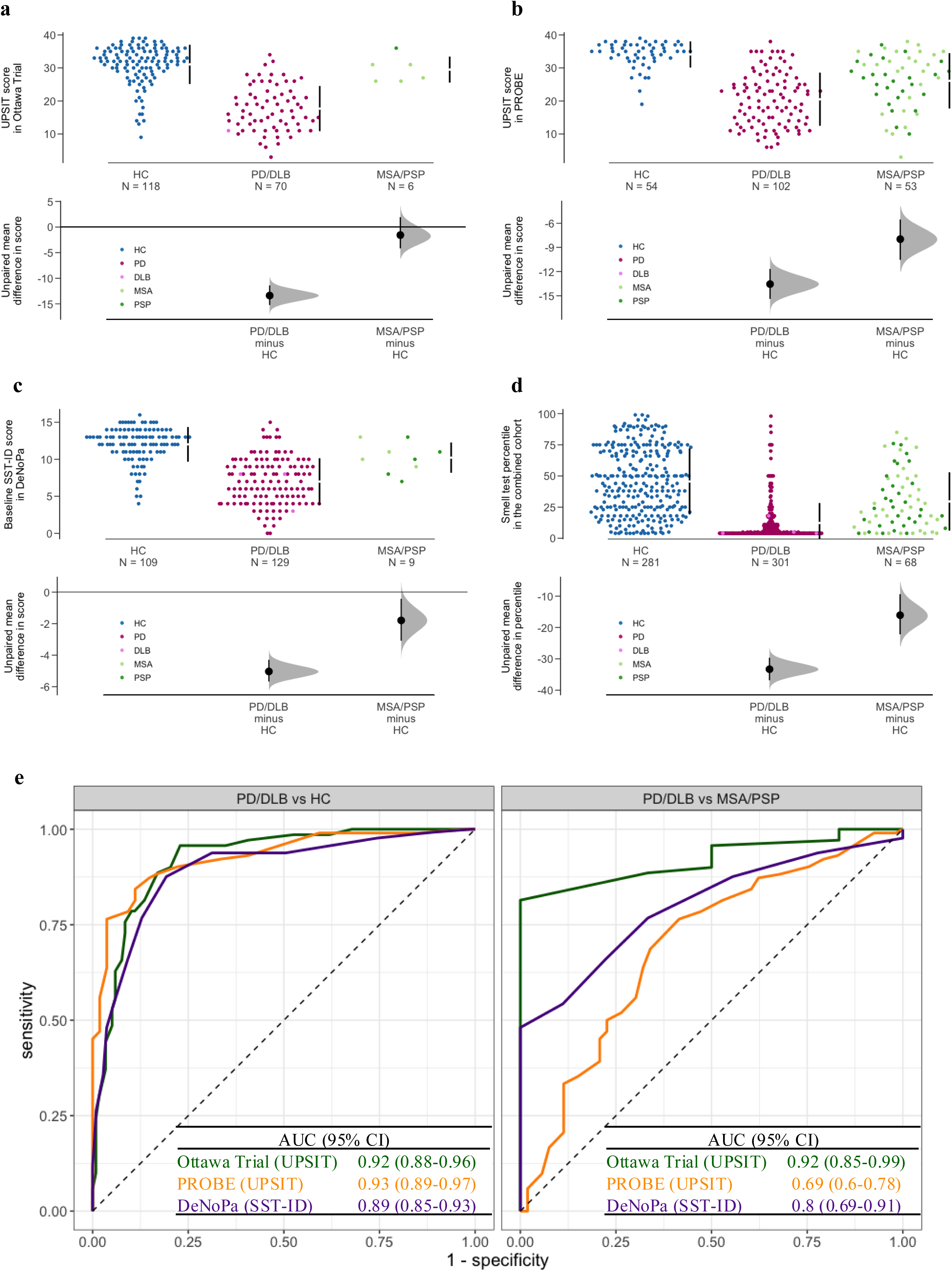
Distribution of olfaction scores using two established tests for different diagnostic groups with parkinsonism in three cohorts. Cummings estimation plots (a-d) were used to illustrate and compare smell test score distributions in each diagnostic group: (a) for UPSIT in the Ottawa Trial cohort, (b) for UPSIT in the PROBE cohort, (c) for SST-ID in the DeNoPa cohort, (d) UPSIT and SST-ID scores were transformed to percentiles based on age- and sex-adjusted norms in the combined cohorts. Each data point in the upper panels represents the score of one participant, and colors represent different groups and diagnosis, as shown in legends. The vertical lines in the upper panels represent the conventional mean ± standard deviation error bars. The lower panels show the mean group difference (the effect size) and its 95% confidence interval (CI) estimated by bias-corrected and accelerated bootstrap, using healthy controls as the reference group. Panels in (e) show ROC curves and AUC values with 95% confidence interval (CI) for smell tests in each cohort (indicated by different colors; individual scores shown in a-d) to distinguish PD/DLB versus HC groups (left) and PD/DLB versus MSA/PSP groups (right). Abbreviations as in Figure 1.

### Performances of individual scents differ in discriminating PD/DLB from HC

We next sought to determine whether subsets of scents were more informative to discriminate between PD/DLB and controls compared to the complete 16-scent (SST-ID) or 40-scent (UPSIT) tests. **Figure 3(a)** shows the distribution of AUC values for each SST-ID scent across 10 folds using baseline data from the DeNoPa cohort. Clusters of scents identified included banana and mint as the two most discriminative scents (individual AUC values, ≥0.725), followed by anise, coffee, licorice, fish and rose in the second-most discriminative cluster. Compared with SST-ID scents, clustering was less obvious for the UPSIT scents (**Figure 3(c)**), whose AUC values ranged between 0.5 to 0.77. For the Ottawa Trial cohort, the top-7 UPSIT scents for identifying PD/DLB patients included rose, wintergreen, root beer, licorice, dill pickle, mint, and grass.

**Figure 3:**
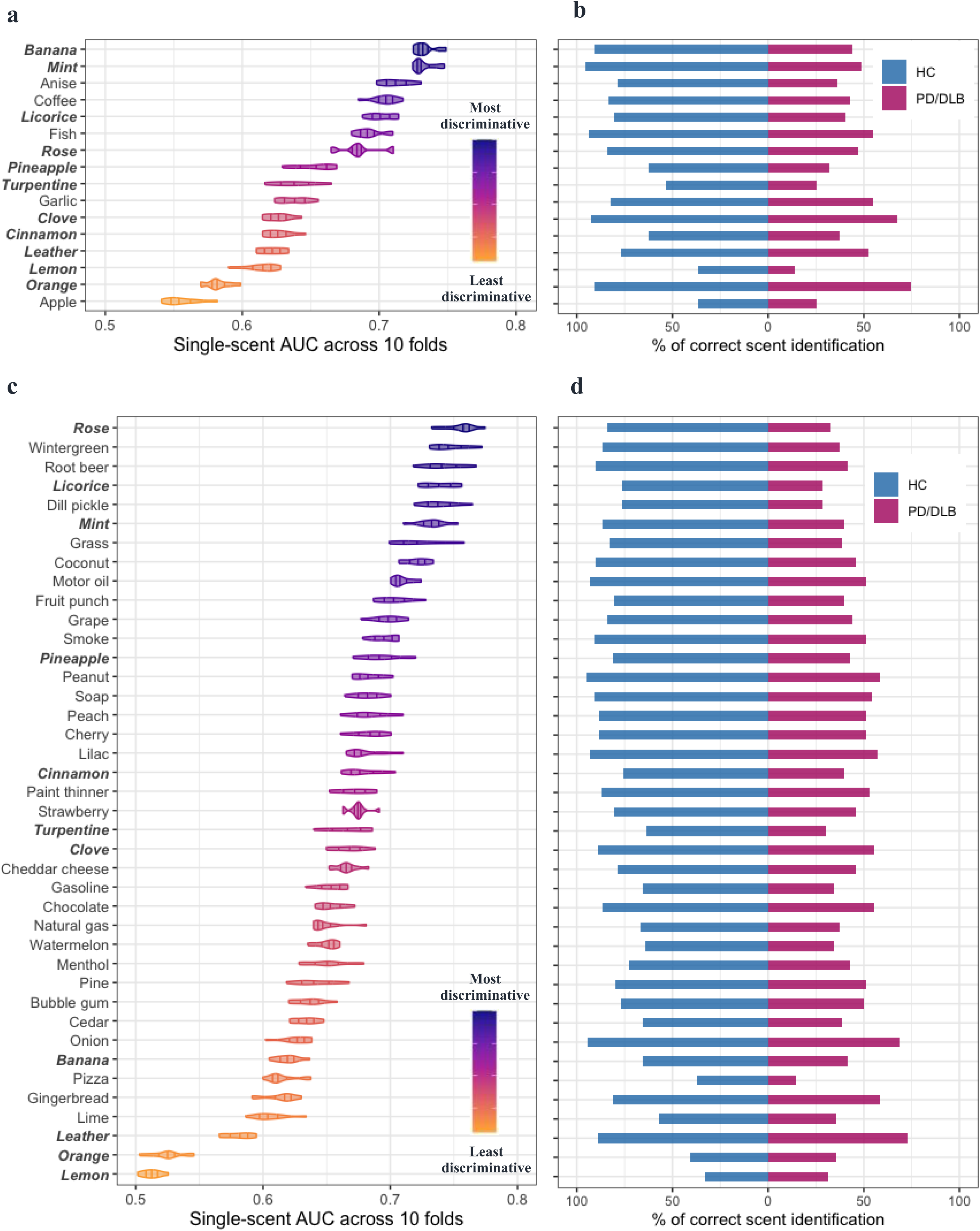
Individual scent performances in differentiating PD/DLB from HC groups. SST-ID scents are shown using baseline DeNoPa data (a, b) and UPSIT scents for the Ottawa Trial cohort (c, d). Panels (a) and (c) illustrate distribution of AUC values of each scent across 10-fold cross-validation using violin plots, with 25%, 50%, and 75% quantile lines. The scents are ordered in descending order of their mean single-scent AUC value; the color of each scent changes gradually from the most to the least discriminative value, as indicated by the legend. Scents shared by both tests are highlighted by bold italic font. Panels (b) and (d) show the percentage of subjects correctly identifying each scent within both groups in each corresponding cohort. Abbreviations as in Figure 1.

The observed differences in each scent’s discriminative performance were further examined by visualizing the percentages of correct scent identification within each diagnostic group (**Figure 3(b), (d)**) and by the percentage differences between HC and PD/DLB groups (**Supplemental Figure 2**). Regardless of the study cohort and smell test used, PD/DLB patients showed significantly lower percentages of correctly identifying each scent than control subjects. Scents that were easy to identify in the HC group but difficult for the PD/DLB group (*i.e.*, larger percentage differences as in **Supplemental Figure 2**) had larger single-scent AUC values. Scents had poorer discriminative performances when both HC and PD/DLB groups found them easy (e.g. SST-ID: orange, UPSIT: leather) or difficult (e.g. SST-ID: apple, UPSIT: lemon).

Therefore, two rankings for scents from the SST-ID and UPSIT kits were constructed. To overcome the inherent risk of overfitting due to the modestly sized cohorts used, external evidence was introduced. **Figure 4** compared the scent rankings from this study with previously published studies, and two “Average” rankings were derived. For the SST-ID kit, different studies -despite their differences in cohort design and methods applied (**Supplemental Table 1**)-showed consensus in that anise, licorice, mint, banana, coffee, fish and rose were the most discriminative scents in distinguishing PD/DLB subjects from HCs (**Figure 4(a)**). For the UPSIT battery, however, its related studies showed less agreement on their scent rankings (**Figure 4(b)**), which could be partially explained by results shown in **Figure 3(c)**; there, many UPSIT scents showing similar performances and revealed fewer clusters than did SST-ID-based scents. Nonetheless, the top-7 UPSIT scents in the final “Average” ranking were coconut, clove, wintergreen, banana, licorice, grass and cherry. With respect to a possibly common, simplified list, we noted that there are eleven scents shared by SST-ID and UPSIT, and thus, an additional “Shared” ranking was generated to construct a unified, abbreviated smell test (**Table 3**).

**Figure 4:**
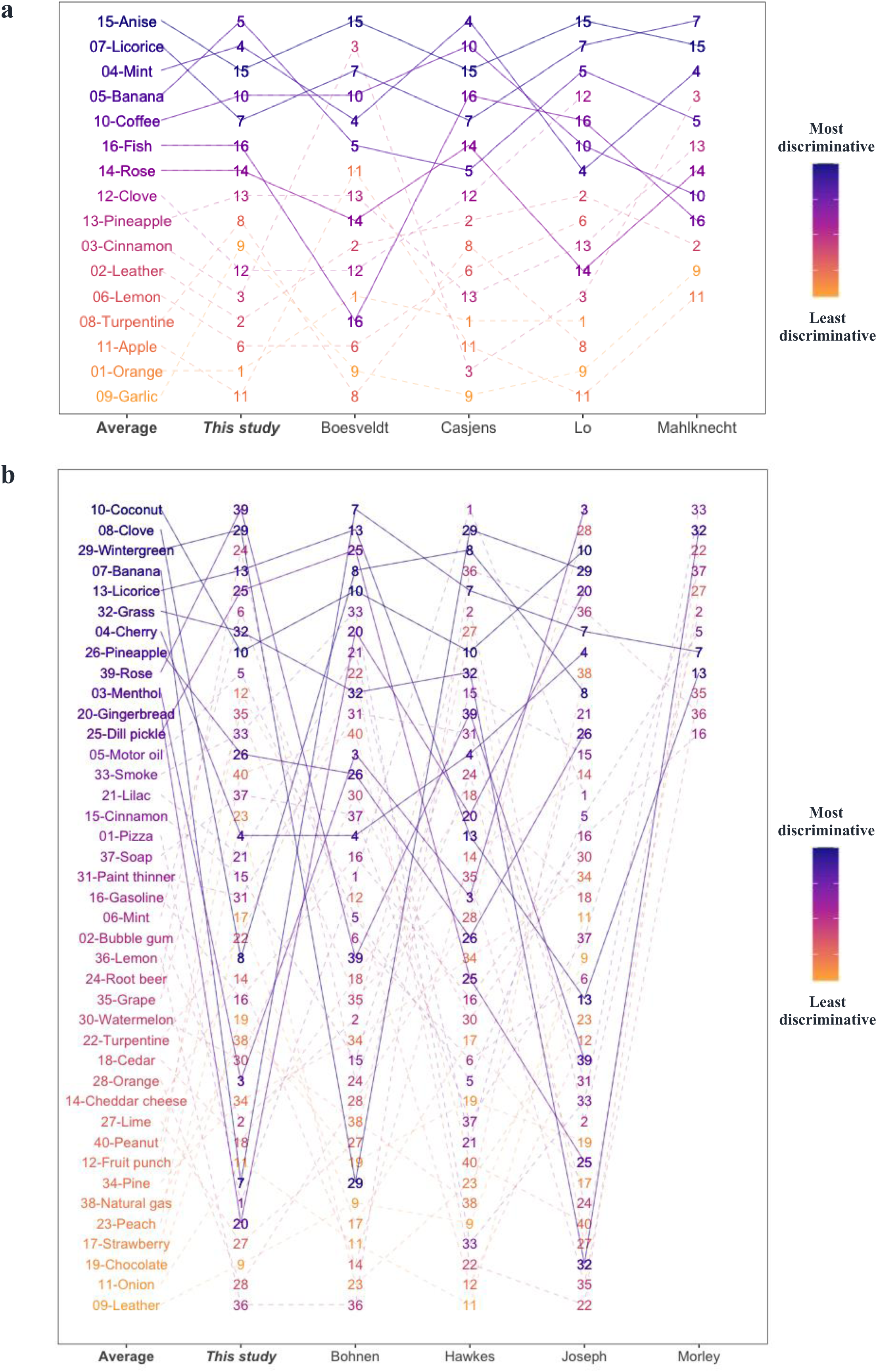
Comparison of scent rankings in this study versus previously published ones. Panels (a) and (b) show scent rankings of SST-ID and UPSIT, respectively. “This study” columns show scent rankings from Figure 3, and the neighboring columns show corresponding rankings from other studies, as indicated at the x-axis. The “Average” column of each panel shows the scent ranking generated by averaging results from 5 separate rankings. Each scent is represented using the format “index-scent” in the “Average” ranking, and as index only in others. The lines track how each scent’s rank changes from study to study. Color of each scent changes gradually from the most to the least discriminative odorant defined by “Average”. Based on these, 7 best-performing scents in SST-ID (a) and 12 best-performing scents in UPSIT (b) are tracked by solid lines. Note, rankings by Mahlknecht *et al.* and Morley *et al.* included only the top 12 scents.

**Table 3:** Ranking of scents that are shared by SST-ID and UPSIT kits. SST-ID = Sniffin’ Sticks Identification test. UPSIT = University of Pennsylvania Smell Identification Test.

| Rank | Scent | Rank in SST-ID<sup>1</sup> | Rank in UPSIT<sup>1</sup> | Average<sup>2</sup> |
| --- | --- | --- | --- | --- |
| 1 | Licorice | 2/16 | 5/40 | 0.125 |
| 2 | Banana | 4/16 | 4/40 | 0.175 |
| 3 | Clove | 8/16 | 2/40 | 0.275 |
| 4 | Rose | 7/16 | 9/40 | 0.33125 |
| 5 | Mint | 3/16 | 21/40 | 0.35625 |
| 6 | Pineapple | 9/16 | 8/40 | 0.38125 |
| 7 | Cinnamon | 10/16 | 16/40 | 0.5125 |
| 8 | Lemon | 12/16 | 23/40 | 0.6625 |
| 9 | Turpentine | 13/16 | 27/40 | 0.74375 |
| 10 | Orange | 15/16 | 29/40 | 0.83125 |
| 11 | Leather | 11/16 | 40/40 | 0.84375 |

<sup>1</sup> Based on the corresponding "Average" rankings in **Figure 4**.
<sup>2</sup> For each scent, average = (rank in SST-ID + rank in UPSIT) / 2

### Item Characteristic Curves further reveal details for each scent and the influence of distractors

The findings above revealed subsets of scents that were relatively discriminative (from a PD perspective), which could suggest a disease-specific and/or scent processing-related change that is linked to *participant performance*. However, for multiple-choice tests like SST-ID and UPSIT kits, the selection of distractors paired with each scent could also influence *odorant performance*. Item Characteristic Curves (ICCs) can help address this, especially when the scent is shared by different tests.

In the current context, mint and licorice were two well-performing scents, and their ICCs (**Figure 5**(1)-(4)) showed similar characteristics in that HCs generally correctly identified them, while PD/DLB patients had more difficulty in choosing the right option. However, there were also some noteworthy differences: when scoring on the scent for ‘mint’, PD/DLB patients could rule out ‘chive’ and ‘onion’ in SST-ID and ‘fruit punch’ in UPSIT, indicating that they detected some scent, but it was not declarative enough to choose ‘mint’. However, for ‘licorice’, particularly in the UPSIT kit, there was strong evidence of random guessing whereby patients couldn’t detect any scent to help favor or eliminate an option. Here, ICCs of scoring by HCs also eliminated the possibility of the corresponding pen (SST-ID) or encapsulated sticker (UPSIT) being defective.

**Figure 5:**
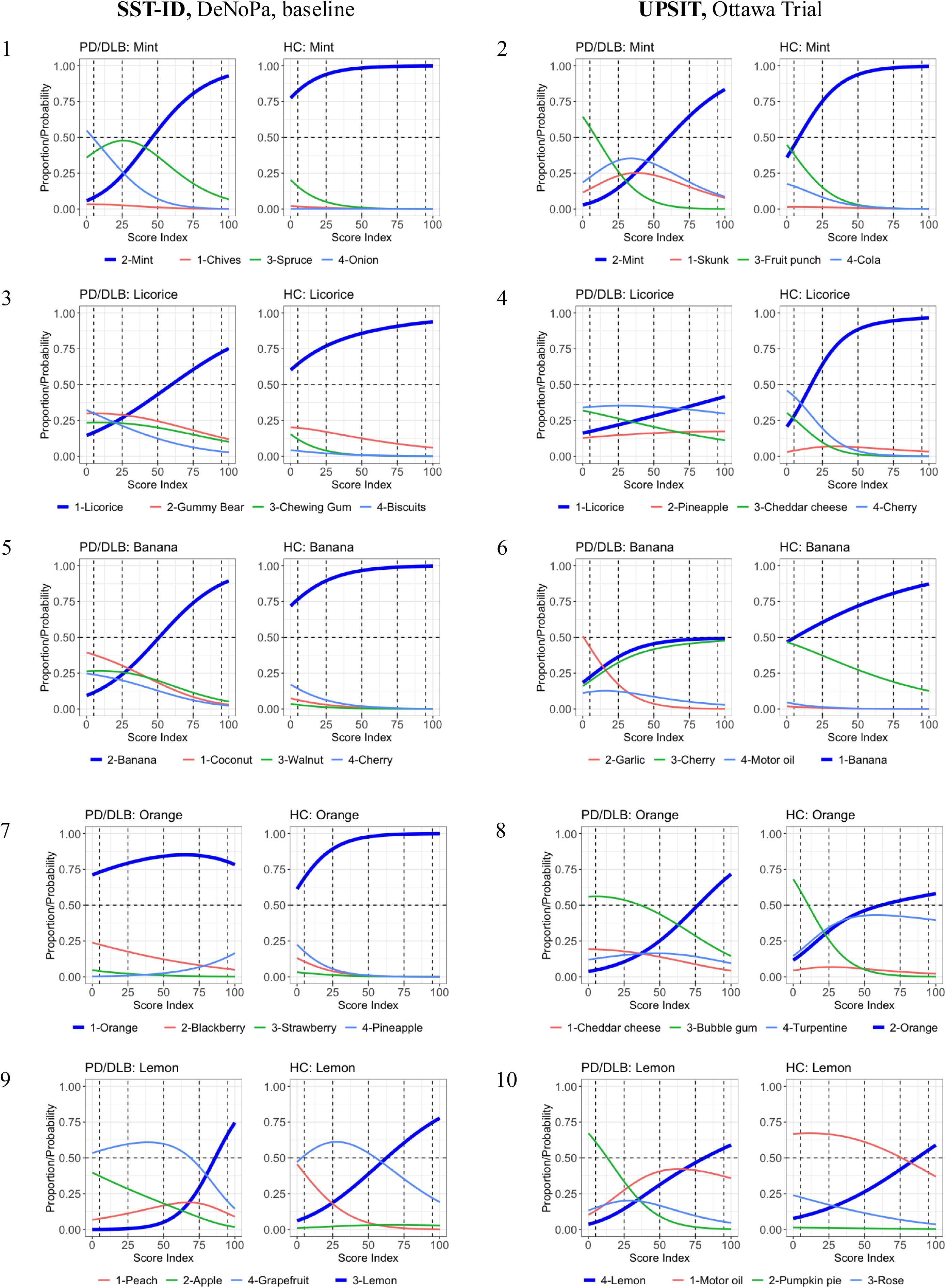
Influence of distractors in multiple-choice smell tests for five shared scents selected. Panels with odd numbers show the Item Characteristic Curves (ICCs) of five SST-ID scents: mint, licorice, banana, orange, and lemon. Panels with even numbers show ICCs of the corresponding UPSIT scents. In each figure, panels on the left show data for PD/DLB patients, panels on the right for healthy controls. The x-axis reveals transformed score indices (percentage rank of the respective scores) within the corresponding group. The y-axis shows the probability of choosing each option at a particular score index. The correct option of each item is highlighted using thicker, blue curves. Numbers in the color legends represent option indices. The horizontal dashed lines represent 50% probability. The vertical dashed lines represent five quantiles (5%, 25%, 50%, 75%, and 95%).

The scent for ‘banana’ ranked 1/16 in DeNoPa but only 34/40 in the Ottawa Trial (**Figure 3**); however, these inconsistent performances were not due to differences in distractors. The option ‘cherry’ distracted many PD/DLB patients and HCs in the Ottawa Trial, but not in the DeNoPa study (**Figure 5**(5)-(6)). Here, cohort- or odorant (*e.g.,* its concentration or composition for the artificial scent)-related differences might be more plausible explanations.

‘Orange’ and ‘lemon’ were both ranked low in the two tests but for different reasons (**Figure 5**(7)-(10)): ‘orange’ in SST-ID was too easy, even for hyposmic PD/DLB patients. ‘Orange’ in UPSIT, however, had different distractors that were active within PD/DLB (‘bubble gum’) and HC (‘turpentine’). For ‘lemon’, the distractors of ‘grapefruit’ in SST-ID and ‘motor oil’ in UPSIT confused both patients and healthy persons. Such ICC results within the normosmic control group might be evidence of a flawed odorant choice or an explanation that is rooted in chemical manufacturing of the scent. In **Supplemental Figures 3**-**5** we listed the ICCs for all other scents.

### Development and validation of abbreviated smell tests

Based on scent rankings in **Figure 4** and **Table 3**, 95 SST-ID subsets and 220 UPSIT subsets were compared by their AUC values in distinguishing PD/DLB patients from HCs. **Figure 6** shows the results within each internal and external validation set. When using an increasing number of highly rank-ordered scents, we observed that the corresponding AUC values for odorant subsets increased steeply for the first four; surprisingly, any improvement in performance thereafter was marginal. Compared with other published rankings, the “Average” rankings as well as their subsets appeared to be more discriminative with robust performance in all the validation sets. Considering a potential trade-off between subset performance and number of scents administered, the SST-ID subset with seven scents and UPSIT subset with ten scents, based on their corresponding “Average” rankings, emerged as the best-performing test batteries.

**Figure 6:**
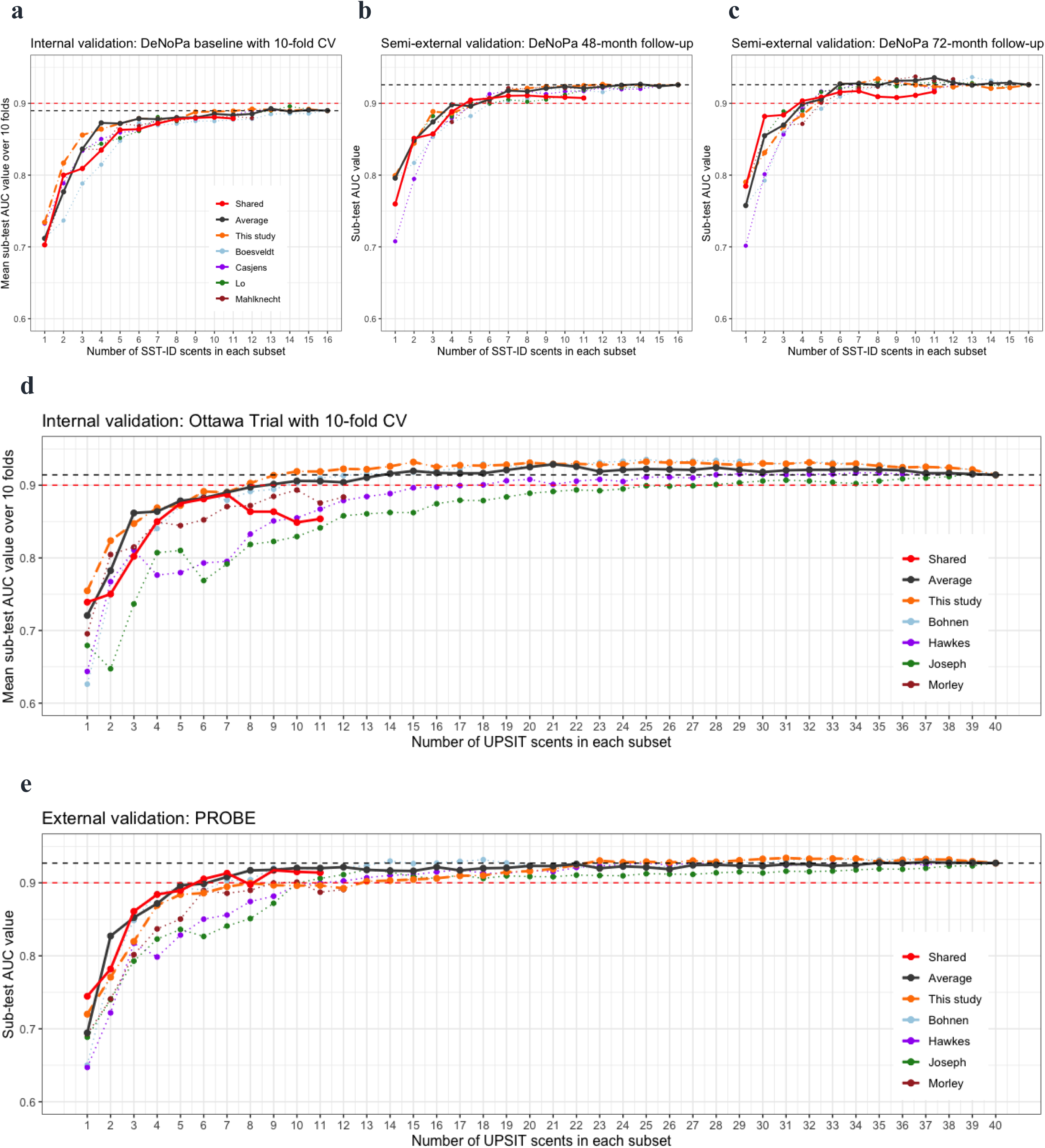
Exploration of smaller subsets of scents tested vs. accuracy in group classification of PD/DLB vs HC. The x-axis shows the number of individual scents used for each subset examined; colors represent different scent rankings from separate studies, as indicated by the legends (see also Figure 4 and **Table 3**). ‘Shared’ denotes scents used in both UPSIT and SST-ID; Average, all studies combined; This study, rankings derived using baseline DeNoPa and Ottawa Trail data. Individual points shown in panels (a) and (d) represent internal validation results, averaging across 10 folds. In panels (b), (c) and (e), each point represents the AUC value of the corresponding subset using (semi-)external validation sets. The black horizontal, dashed lines indicate AUC values of the corresponding test when viewed in its entirety. Red horizontal, dashed lines indicate AUC = 0.9 as a predetermined reference line.

With respect to potentially developing a unified smell test that could be applied to large study populations (those examined here were based in North America and Europe of predominantly White ethnicity), the subset of seven scents from the “Shared” ranking (**Table 3**) with the highest performance in all validation sets comprised licorice, banana, clove, rose, mint, pineapple and cinnamon. Mean (standard deviation) scores and AUC values of this subset within each cohort are reported in **Table 4**. Note, due to inherent cohort differences, the AUC value in the diagnostic classification of PD/DLB versus HC when combining all three cohorts was 0.87 (95%CI 0.85-0.9), *i.e.,* slightly lower than those of each separate cohort. In the combined cohort, the cut-off value for distinguishing PD/DLB versus HC that corresponded to the maximum Youden index was 4.5 (score range 0-7), the resulted sensitivity and specificity were 0.76 and 0.85, respectively.

**Table 4:**
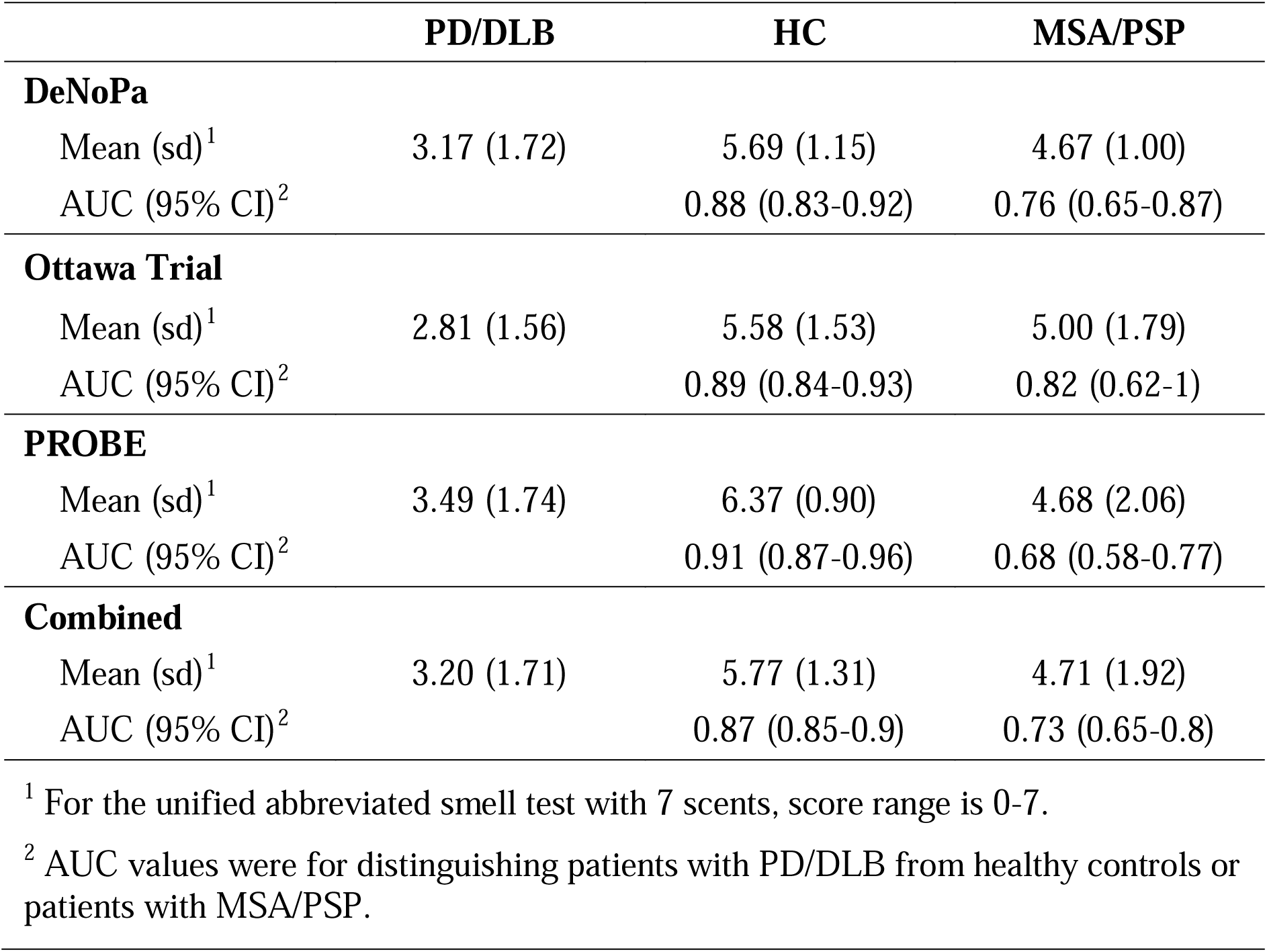
Performance of the unified abbreviated smell test with 7 scents. DeNoPa = *De Novo* Parkinson Study. PROBE = Prognostic Biomarkers in Parkinson Disease. HC = healthy control. PD = Parkinson disease. DLB = dementia with Lewy bodies. MSA = multiple system atrophy. PSP = progressive supranuclear palsy. AUC = area under the ROC curve. CI = confidence interval. sd = standard deviation.

### Performances of scents in discriminating PD/DLB from MSA/PSP

The same workflow was applied to the PROBE cohort to generate a subset of scents specialized in distinguishing between PD/DLB versus MSA/PSP patients. There, a subset with ten scents (clove, dill pickle, cinnamon, soap, rose, pizza, root beer, turpentine, gasoline, licorice) achieved an AUC value 0.78 in the validation set within PROBE, a promising improvement when compared with the entire 40-scent UPSIT test (AUC = 0.68; **Supplemental Figure 6**). Independent validation is needed to retest the usefulness of this subset given the small number of MSA/PSP subjects enrolled in the other two cohorts studied herein.

### Assessment of age and sex on scent identification

We also investigated the influence of age and sex on scent identification. **Supplemental Table 2** shows the coefficients of the linear regression for the relationship between smell test score with age, sex, and diagnostic groups within each cohort. Not surprisingly, progression in age significantly lowered olfaction across all groups, and males generally had a worse sense of smell than their female counterparts, although the latter was not significant across the three cohorts.

When focusing just on the eleven scents shared by both tests, relationships between scent identification and sex were further evaluated by comparing the percentages of correct scent identification across groups (**Figure 7(a)**). In line with the regression results, females showed higher percentages of correct identification than males for most of the scents, except for cinnamon, turpentine, and leather. Next, we compared the probability of identifying each scent correctly across ages between the PD/DLB and HC groups (**Figure 7(b)**). As expected, older participants showed decreasing percentages for identifying specific scents correctly. The fitted lines for PD/DLB and HC groups were usually in the same direction and of similar slopes, with some exceptions, but these were not consistent across all three cohorts.

**Figure 7:**
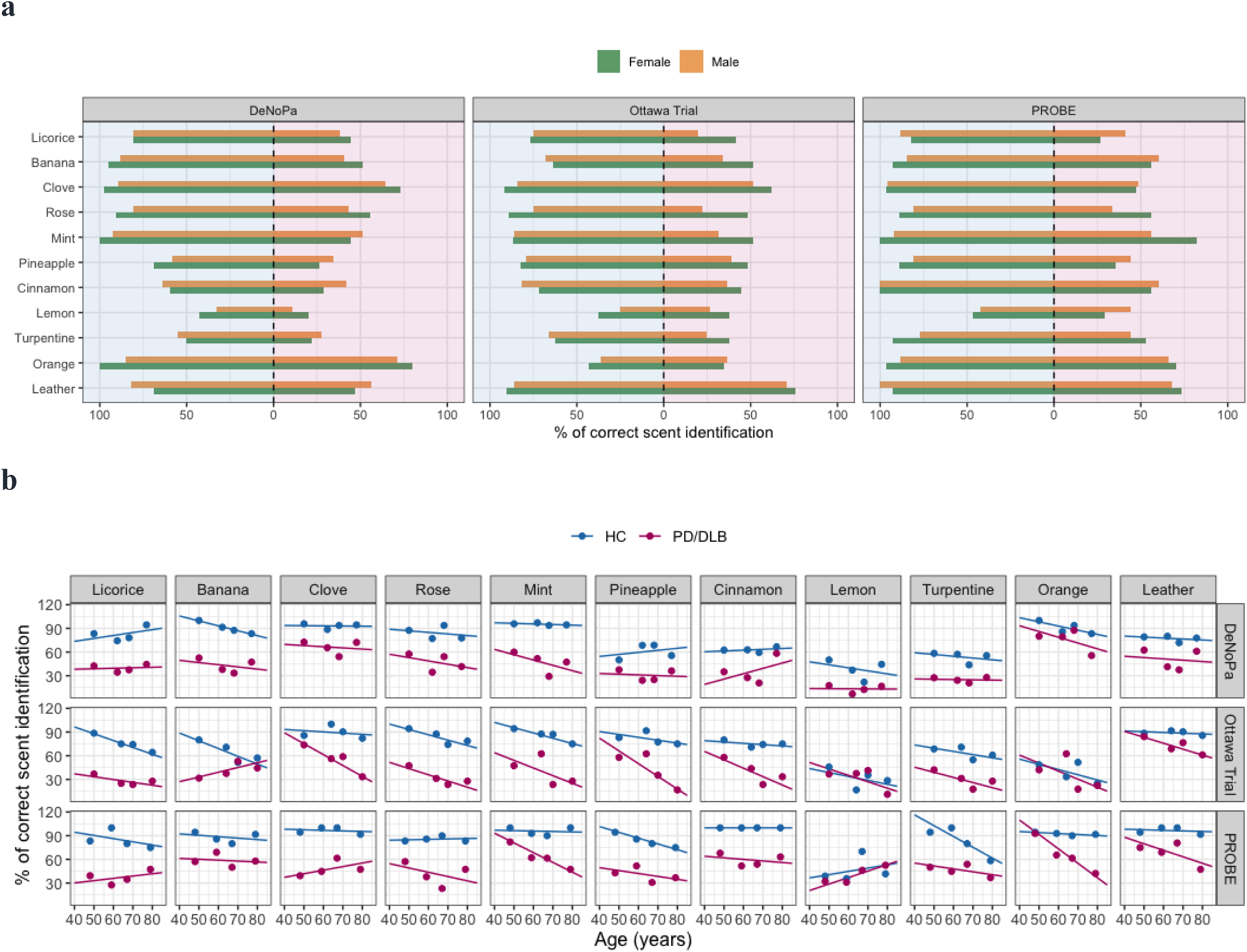
Relationships between scent identification performance, diagnosis, sex and age. Panel (a) shows the percentage of persons that correctly identified each scent within the healthy control (HC) group (indicated by light blue region) and the PD/DLB group (indicated by pink region) in the corresponding cohorts, separated by sex (bar color). Panels in (b) show the relationship between age (x-axis), diagnostic group (HC in blue; vs PD/DLB in red) and the percentage of correctly identified scents (y-axis) for each odorant tested (columns) within each cohort (row), as indicated on the right.

## DISCUSSION

To our knowledge, this is the most comprehensive study to date describing olfactory dysfunction in late-onset, typical PD and two other neurological disorders presenting with parkinsonism using both the multimodal SST battery and UPSIT kit. The principal take home message from this study is that when probing for hyposmia in PD, the following points matter: PD/DLB patients had worse olfaction than healthy subjects, and scores of MSA/PSP patients were intermediate; there was no observed difference in olfaction between MSA and PSP patients; scent identification testing is sufficient, and threshold as well as discrimination testing could be omitted when screening populations for PD using the SST kit; fewer scents can reduce examination time and test taking fatigue without sacrificing diagnostic accuracy; the selection of specific scents should be informed by their discriminative performance in specific group classification efforts; random guessing could lower diagnostic accuracy; and from a test design perspective, choices provided as distractors influence scent performance. Importantly, we found that a simplified smell test, with specific scents, is sufficient to identify PD/DLB-linked hyposmia. Such a test, which is now being piloted by us, holds the potential to facilitate olfactory testing in the clinical setting, used for at-home testing and population-based screening methods.

In developing and validating a simplified smell test for this purpose, we used a machine learning approach and found that only seven scents (licorice, banana, clove, rose, mint, pineapple, and cinnamon) were required to approximate the diagnostic performance of administering the 16-scent SST-ID or 40-scent UPSIT batteries and the value of adding more scents was negligible. Moreover, a test kit for rapid screening with nearly similar performance could be constructed using as few as four scents.

We demonstrated the impact distractors have on detecting specific scents using IRT analysis. We uncovered uncertainty in eliciting a choice for some scents, even for HCs with intact olfaction. This could be explained by the difficulty of biological scent discrimination or the to-be-improved selection of artificial odorants. By extension, our analyses revealed the opportunity to remove ill-performing scents*, e.g.,* orange and lemon, from currently used kits.

Unexpectedly, we found a high level of guessing among PD patients for licorice, indicating patients’ difficulty in detecting this scent. SST-ID and UPSIT batteries are multiple choice-based tests, in which participants are instructed to always choose even when they cannot smell anything; such random guessing will introduce errors into data sets. Advanced IRT methods can treat missing responses as an additional option; administrators of tests would then prefer the participants to leave any uncertain questions unanswered rather than forcing a guess. However, for the future administration of olfaction tests, or for designing a new one, we would suggest adding an extra choice, such as “I cannot identify the scent” to reduce random guessing. Based on our experience in administrating smell tests, the extra option would also help improve participant experience and eliminate frustration, especially for patients with severe hyposmia.

An easy-to-administer, inexpensive, sensitive and non-invasive smell test (with 4-7 scents) could have important clinical usefulness, particularly when coupled with a short, self-administered questionnaire capturing demographic information and known risk factors of developing PD.^[9]^ Such a questionnaire could also explore other factors leading to hyposmia unrelated to neurodegeneration, *e.g.,* previous nasal injuries, microbial infections, seasonal allergies, and chronic exposure to air pollution, to augment specificity for PD. Upon validation, such a kit could be used as the initial step of large-scale community screening, or in routine clinic practice of a movement disorders-oriented clinic, or for early detection within a family medicine office. When it comes to screening efforts for PD, more invasive and expensive tests, *e.g.,* the α-synuclein seeding amplification assay from cerebrospinal fluid (CSF), or the administration of a dopamine transporter scan, could be administrated as an additional step to increase screening accuracy further, such as when considering enrolling PD subjects into specific, disease-modifying clinical trials.^[40]-[44]^

Despite the findings regarding scent ranking and subset analyses, it remains unclear whether a specific PD olfaction deficit exists, rather than a global reduction in olfaction, and what the underlying mechanisms would be. We and others recently found that olfactory deficits were significantly associated with positivity on the α-synuclein seeding amplification assay in CSF samples, suggesting that patients may have an underlying disease linked to the dysregulation of *SNCA* expression and/or protein processing.^[45],[46]^

Mechanistically, it remains unknown as to how chronic hyposmia arises in PD/DLB (and REM Sleep Behaviour Disorder) as well as some MSA/PSP subjects, at what age it begins, the cause and its underlying circuit-based and molecular mechanisms, and whether olfactory deficits are shared for specific scents among persons with typical PD versus those with dementia. Large scale population screening, including with a simplified testing battery derived from SST-ID and UPSIT kits, could begin to answer some of these questions.

## STRENGTH

In this work, olfaction performance using SST and UPSIT batteries was studied in detail. Both internal and external validation efforts were used to test/retest performance and avoid overfitting. Scent ranking derived from this study was also compared with eight previously published studies. By incorporating results from external studies, the unified abbreviated olfaction test kit of using just seven scents emerged as very generalizable. The observed performance differences for each scent in group classification was explained using complementary techniques and further studied considering sex and age. Our study also explored scent identification performance at the level of choices provided, highlighting the importance of distractors, which allows for future improvement in the design of test kits.

## LIMITATIONS AND FUTURE WORK

Performance of the unified abbreviated smell test was estimated by simulation: responses of the related scents were partitioned from the SST-ID and UPSIT data sets, and the original distractors from SST-ID and UPSIT kits were used. Real performance of the abbreviated test, when manufactured as a stand-alone product with distractors determined for each scent, should be assessed in the same cohorts, in new studies, and in routinely run clinic settings. Further, more data are needed to validate the scent ranking and the associated subset developed here for distinguishing PD/DLB versus MSA/PSP patients.

The three cohorts used are highly homogeneous cohorts with most participants being white. Although scent rankings and the selection of a simplified smell test have been rigorously developed and validated with external information incorporated, future calibration and cultural adaption efforts will be necessary when applying them to other populations, especially those that differ from Western Europeans.

Case-control studies have an inherent potential for selection bias in their recruitment. Especially because of the age- and sex-matched design, age- and sex-effects were likely underestimated. Population studies, such as in the initial community screening effort undertaken by PARS planners and the ‘PPMI hyposmia’ effort, could provide complementary data sets; however, they too have potential setbacks: as the majority of participants will have a normal sense of smell, the score distribution could be highly skewed; a low percentage of PD patients will make the resulting dataset imbalanced; when smell test data are reduced to a single sum score, sub-analyses will be difficult to complete, which will limit interrogations of data between established cohorts.

For screening purpose, a one-time administered smell test may not be informative enough to assess a subject’s sense of smell completely, because other factors, such as temporary olfaction reduction/loss due to infection, seasonal allergies, occupational exposure and/or drinking, eating, smoking before taking the test, could skew results. Retesting at appropriate time intervals will be required for even higher accuracy.

## ACKNOWLEDGEMENTS

The authors acknowledge the commitment of study participants in the three cohorts and are grateful to all clinical research coordinators at all the study sites. Responsible supervisors for the:

DeNoPa Study: Paracelsus-Elena-Klinik: Brit Mollenhauer.

Ottawa Trial Study: The Ottawa Hospital: Michael Schlossmacher, Élisabeth Bruyère Hospital: Andrew Frank.

PROBE Study: PROBE Steering Committee: Voyager Therapeutics: Bernard Ravina; Brigham and Women’s Hospital: Clemens Scherzer, University of Ottawa: Michael Schlossmacher, Avid Radiopharmaceuticals: Andrew Siderowf, University of Rochester: David Oakes, Arthur Watts; Institute for Neurodegenerative Disorders: Kenneth Marek; Georgetown University: Ira Shoulson.

We thank Nathalie Lengacher for help in graphic design; Nadine Mauri, Nancy MacDonald, and Yoobin Lee for help in data management/input. This work was supported by funding from Parkinson Canada (to T.M., D. M., M.G.S; 2018; to J.L.; 2019-2021), Michael J. Fox Foundation for Parkinson’s Research (to T.M., D. M., M.G.S), Department of Medicine (T.M., T.R., D.M., M.G.S.), The Ottawa Hospital Foundation (Borealis Foundation to J.L.) and the Uttra & Subhash Bhargava Family (M.G.S.), the Paracelsus-Elena-Klinik Kassel, Parkinson Fonds Deutschland, and the Deutsche Parkinson Vereinigung (B.M.; C.T.). The study was also funded by the joint efforts of The Michael J. Fox Foundation for Parkinson’s Research (MJFF) and the Aligning Science Across Parkinson’s (ASAP) initiative. MJFF administers the grant [Grant ID: ASAP-020625] on behalf of ASAP and itself.

The funders had no role in the design and execution of the study; the collection, management, analysis, and interpretation of the data; the preparation, review, or approval of the manuscript; and the decision to submit the manuscript for publication. We are grateful for the ongoing support and feedback from people with lived experiences, such as through the board of the Parkinson’s Research Consortium Ottawa and members of Partners Investing in Parkinson’s Research, and to Drs. P. Wells and D. Lewis for their ongoing encouragement.

## AUTHOR CONTRIBUTIONS

JL and MGS contributed to the concept and design of the study; JL, KG, and MGS contributed to the acquisition of data. JL decided on the statistical methods used in this study. JL did data cleaning, data analysis, figures and tables. JL, JJT, and MGS contributed to data interpretation. SS, SW, MD, TW, EL, CT, and BM contributed to the data collection and verification of DeNoPa; KG, JS, JL, JJT, AF, and MGS contributed to the data collection and verification of Ottawa Trial. JL and MGS wrote the first draft of the manuscript. JJT, BM, NS, TAM, TR, and DM contributed to the drafting of the manuscript and revising it critically, and all authors approved the submission of its current version.

## CODE AVAILABILITY

The code for data analyses and figures is publicly accessible in GitHub (link to be updated).

## DATA AVAILABILITY

The datasets used in this study can be accessed via zenodo (link to be updated) upon request.

## SUPPLEMENTAL METHODS

### Detailed description of the study cohorts

*De Novo* Parkinson disease study (DeNoPa): The DeNoPa cohort^[27]^ is an ongoing, single-center study based in Kassel, Germany. The DeNoPa cohort is an observational, longitudinal study of patients with a newly established diagnosis of PD (UK Brain Bank Criteria^[29]^), who were naïve to L-DOPA therapy at baseline, and of age- and sex- and education-matched, neurologically healthy controls (HC). Details of inclusion/exclusion criteria have been described elsewhere.^[27]^ Diagnostic accuracy was ensured by ongoing follow-up visits every two years (as of 2023, 10-year follow up visits were underway). Data used were received on May 16^th^, 2023.

Ottawa (PREDIGT) Trial: The Ottawa Trial is a pilot study to evaluate the performance of a 2-step screening tool that combines the PREDIGT questionnaire^[8],[9]^ and the UPSIT test to distinguish patients with PD/DLB from age-matched neurologically healthy controls and patients with various other neurological diseases. Enrolment and assessment of this cross-sectional, case-control study was completed in March 2024. A manuscript that describes this cohort is in preparation. Diagnostic accuracy was ensured by independent chart review by three subspecialty-trained neurologists (JS, MGS, and AF) according to UK Brain Bank Criteria and MDS Criteria.^[30]^

Prognostic Biomarkers in Parkinson Disease (PROBE): PROBE^[28]^ is a longitudinal, case-control study to test biomarkers in PD subjects and controls to determine their feasibility and potential utility as markers of risk and prognosis for PD. Details of inclusion/exclusion criteria have been described elsewhere.^[28]^ Participants were enrolled from August, 2007 to December, 2008. Diagnosis of PD, probable MSA, and probable PSP met UK Brain Bank criteria, Consensus Criteria, and NINDS-PSP Criteria, respectively.

### Supplemental methods

#### 10-fold cross validation (CV)

For each smell test, the discovery dataset was randomly partitioned into 10 parts, where the case-control ratio was maintained in each part. In each fold, 9/10 parts were used as the development set and the remaining 1 part was used for internal validation. This procedure was repeated for 10 times, and results were showed either in distribution or average across 10 folds.

#### Relationship between scent identification and age

Within each cohort, participants’ ages were segregated into four bins with similar sample size. For each scent, the proportion of correctly identification was calculated for each bin, and spline smoothing was then used to represent the relationship between the proportions and age.

#### Item characteristic curves (ICCs)

The version of ICCs used in this study differed from traditional parametric ICCs in two aspects: 1) the x-axis was the score percentage rank in ^[0,100]^, not the latent trait on the whole real line; and 2) ICCs represented spline smoothing lines that fit response data, rather than being fitted to any pre-defined parametric model.^[35]^

## SUPPLEMENTAL FIGURES

**Supplemental Figure 1: Distribution of Sniffin’ Sticks Threshold (SST-TH) and Discrimination (SST-DS) scores for each subject group in the DeNoPa Study at baseline and their ROC curves for group classification.** The Cummings estimation plots (a, b) were used to illustrate and compare smell test score distributions in each group: (a) SST-TH, (b) SST-DS. Each data point in the upper panels represents the score of one participant, and colors represent different groups and diagnosis as shown in the legend. The vertical lines in the upper panels represent the conventional mean ± standard deviation error bars. The lower panels show the mean group difference (the effect size) and its 95% confidence interval (CI) estimated by bias-corrected and accelerated bootstrap, using HC as the reference group. Panel (c) shows the ROC curves of each smell test (indicated by color) to distinguish PD/DLB versus HC (left) and PD versus OND (right). HC = healthy control. PD = Parkinson disease. DLB = dementia with Lewy bodies. MSA = multiple system atrophy. PSP = progressive supranuclear palsy. ROC = receiver operating characteristic. AUC = area under the ROC curve.

**Supplemental Figure 2: Percentage differences of correct scent identification between HC and PD/DLB groups (% HC - % PD/DLB) in the DeNoPa (a) and Ottawa Trial (b) cohorts.** The scents are ordered in descending orders of their mean single-scent AUC value (see Figure 3 (a) and (c)); the color of each scent changes gradually from the most discriminative to the least discriminative odorant, as indicated by the legend.

**Supplemental Figure 3: Influence of distractors in the multiple-choice smell tests: the remaining 6 scents shared by UPSIT and SST-ID.** Panels with odd numbers show the Item Characteristic Curves (ICCs) of six SST-ID scents, and panels with even numbers show the ICCs of the corresponding UPSIT scents. In each figure, the left panels are the ICCs using data of the PD/DLB patients, and the right panels are corresponding to healthy controls. The x-axis is transformed score indices (percentage rank of the SST-ID score) within the corresponding group. The y-axis is the probability of choosing each option at a particular score index. The correct option of each item is highlighted using the thicker blue curves. Numbers in the color legends are the option indices. The horizontal dashed lines represent 50% probability. The vertical dashed lines represent five quantiles (5%, 25%, 50%, 75%, and 95%).

**Supplemental Figure 4: Item Characteristic Curves (ICCs) of the remaining SST-ID scents using baseline data from the DeNoPa Cohort.** In each panel, the left panels are the ICCs using data of the PD/DLB patients, and the right panels are corresponding to healthy controls. The x-axis is transformed score indices (percentage rank of the SST-ID score) within PD and HC group, respectively. The y-axis is the probability of choosing each option at a particular score index. The correct option of each item is highlighted using the thicker blue curves. Numbers in the color legends are the option indices. The horizontal dashed lines represent 50% probability. The vertical dashed lines represent five quantiles (5%, 25%, 50%, 75%, and 95%).

**Supplemental Figure 5: Item Characteristic Curves (ICCs) of the remaining UPSIT scents using the Ottawa Trial cohort.** In each panel, the left panels are the ICCs using data of the PD/DLB patients, and the right panels are corresponding to healthy controls. The x-axis is transformed score indices (percentage rank of the SST-ID score) within PD and HC group, respectively. The y-axis is the probability of choosing each option at a particular score index. The correct option of each item is highlighted using the thicker blue curves. Numbers in the color legends are the option indices. The horizontal dashed lines represent 50% probability. The vertical dashed lines represent five quantiles (5%, 25%, 50%, 75%, and 95%).

**Supplemental Figure 6: Range of performances for UPSIT scents in differentiating PD/DLB from MSA/PSP subjects in the PROBE cohort and AUC values for numerical subsets of odorants in group classification.** Panel (a) illustrates distribution of AUC values of each scent across 10-fold cross-validation using violin plots, with 25%, 50%, and 75% quantile lines. The scents are ordered in descending order of their mean single-scent AUC value. The color of each scent changes gradually from the most to the least discriminative odorant, as indicated by the legend. Panel (b) shows the percentage of subjects correctly identifying each scent within the MSA/PSP and PD/DLB groups, and panel (c) shows the percentage differences between the two groups. Scents in panels (b) and (c) follow the same rank order as in panel (a). In panel (d), the x-axis is the number of scents included for each subset, with individual points representing average AUC values for the validation set across ten CV folds gathered in PROBE. The black horizontal, dashed line indicates the corresponding AUC values of the whole test (= 40 scents). The red horizontal, dashed line indicates AUC = 0.8 as a predetermined reference line.

## SUPPLEMENTAL TABLES

**Supplemental Table 1:** STARD checklist.

| Section & Topic | No | Item | Reported on page # |
| --- | --- | --- | --- |
| TITLE OR ABSTRACT |  |  |  |
|  | 1 | Identification as a study of diagnostic accuracy using at least one measure of accuracy (such as sensitivity, specificity, predictive values, or AUC) | P1 |
| ABSTRACT |  |  |  |
|  | 2 | Structured summary of study design, methods, results, and conclusions (for specific guidance, see STARD for Abstracts) | P3 |
| INTRODUCTION |  |  |  |
|  | 3 | Scientific and clinical background, including the intended use and clinical role of the index test | Research in context: P4-5<br>Introduction: P6-7 |
|  | 4 | Study objectives and hypotheses | P7 |
| METHODS |  |  |  |
| Study design | 5 | Whether data collection was planned before the index test and reference standard were performed (prospective study) or after (retrospective study) | P7 |
| Participants | 6 | Eligibility criteria | P7, Supplemental methods: P2 |
|  | 7 | On what basis potentially eligible participants were identified (such as symptoms, results from previous tests, inclusion in registry) | P7, Supplemental methods: P2 |
|  | 8 | Where and when potentially eligible participants were identified (setting, location and dates) | Supplemental methods: P2 |
|  | 9 | Whether participants formed a consecutive, random or convenience series | P7, Supplemental methods: P2 |
| Test methods | 10a | Index test, in sufficient detail to allow replication | Description of SST and UPSIT: P8<br>The abbreviated smell test: P13 |
|  | 10b | Reference standard, in sufficient detail to allow replication | Supplemental methods: P2 |
|  | 11 | Rationale for choosing the reference standard (if alternatives exist) | Supplemental methods: P2 |
|  | 12a | Definition of and rationale for test positivity cut-offs or result categories of the index test, distinguishing pre-specified from exploratory | P9 |
|  | 12b | Definition of and rationale for test positivity cut-offs or result categories of the reference standard, distinguishing pre-specified from exploratory | Supplemental methods: P2 |
|  | 13a | Whether clinical information and reference standard results were available to the performers/readers of the index test | P7 |
|  | 13b | Whether clinical information and index test results were available to the assessors of the reference standard | P7 |
| Analysis | 14 | Methods for estimating or comparing measures of diagnostic accuracy | P8-10 |
|  | 15 | How indeterminate index test or reference standard results were handled | Supplemental methods: P2 |
|  | 16 | How missing data on the index test and reference standard were handled | Index test: P8. No missing data on the reference standard. |
|  | 17 | Any analyses of variability in diagnostic accuracy, distinguishing pre-specified from exploratory | P9 |
|  | 18 | Intended sample size and how it was determined | P7 |
| RESULTS |  |  |  |
| Participants | 19 | Flow of participants, using a diagram | P7, Supplemental methods: P2 |
|  | 20 | Baseline demographic and clinical characteristics of participants | Table 1 |
|  | 21a | Distribution of severity of disease in those with the target condition | P7, Table 1 |
|  | 21b | Distribution of alternative diagnoses in those without the target condition | Table 1 |
|  | 22 | Time interval and any clinical interventions between index test and reference standard | P7, Supplemental methods: P2 |
| Test results | 23 | Cross tabulation of the index test results (or their distribution) by the results of the reference standard | Figure 2;<br>Supplemental Figure 1;<br>Tables 1, 2, 4;<br>P10-11, P13-14 |
|  | 24 | Estimates of diagnostic accuracy and their precision (such as 95% confidence intervals) | Figures 2, 6;<br>Supplemental Figures 1, 6;<br>Tables 2, 4;<br>P10-11, P13-14 |
|  | 25 | Any adverse events from performing the index test or the reference standard | The smell tests are non-invasive and there was no adverse event from performing them. P16 |
| DISCUSSION |  |  |  |
|  | 26 | Study limitations, including sources of potential bias, statistical uncertainty, and generalisability | P17-18 |
|  | 27 | Implications for practice, including the intended use and clinical role of the index test | P5, P15-17 |
| OTHER<br>INFORMATION |  |  |  |
|  | 28 | Registration number and name of registry | Supplemental methods: P2 |
|  | 29 | Where the full study protocol can be accessed | Supplemental methods: P2 |
|  | 30 | Sources of funding and other support; role of funders | P4, P19 |

**Supplemental Table 2:**
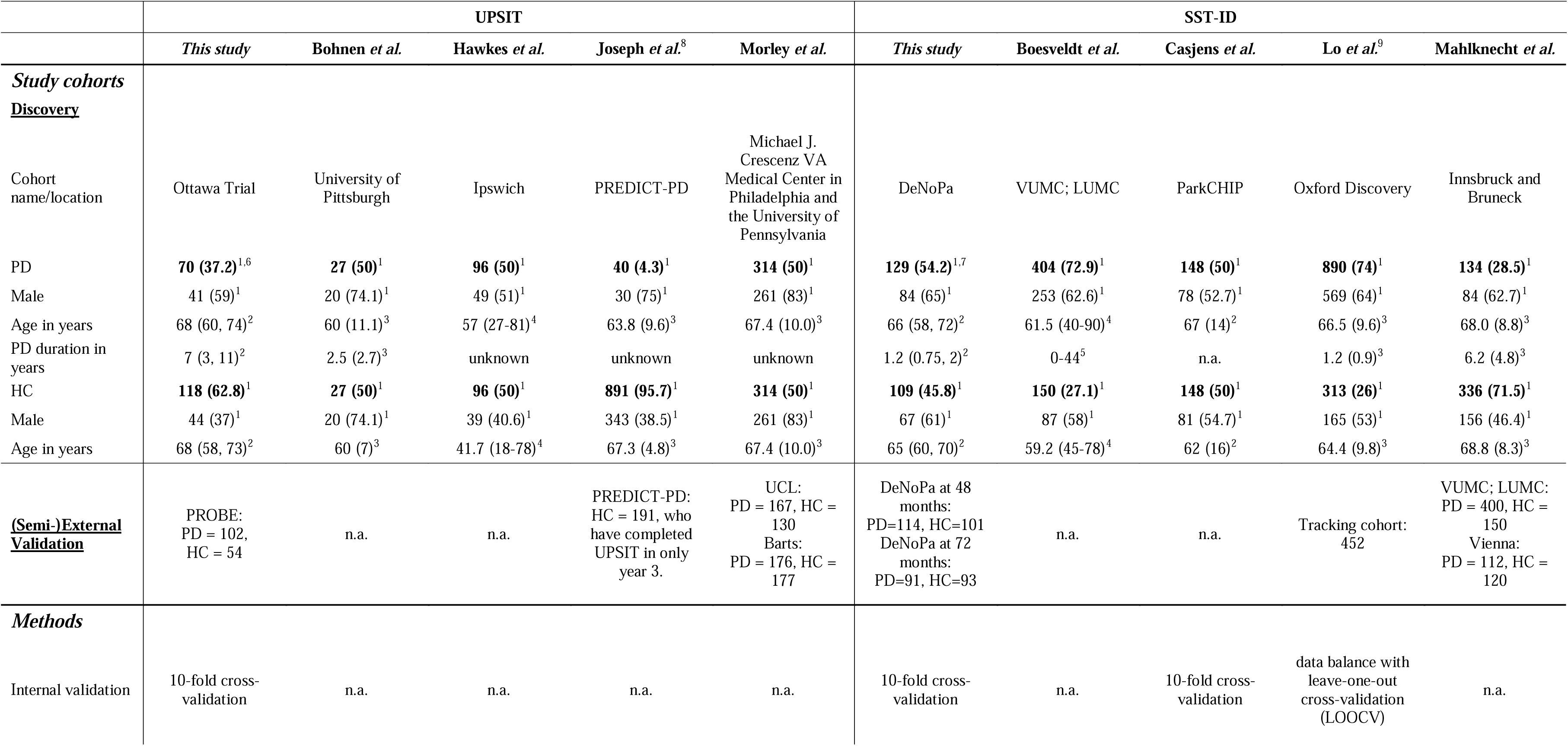
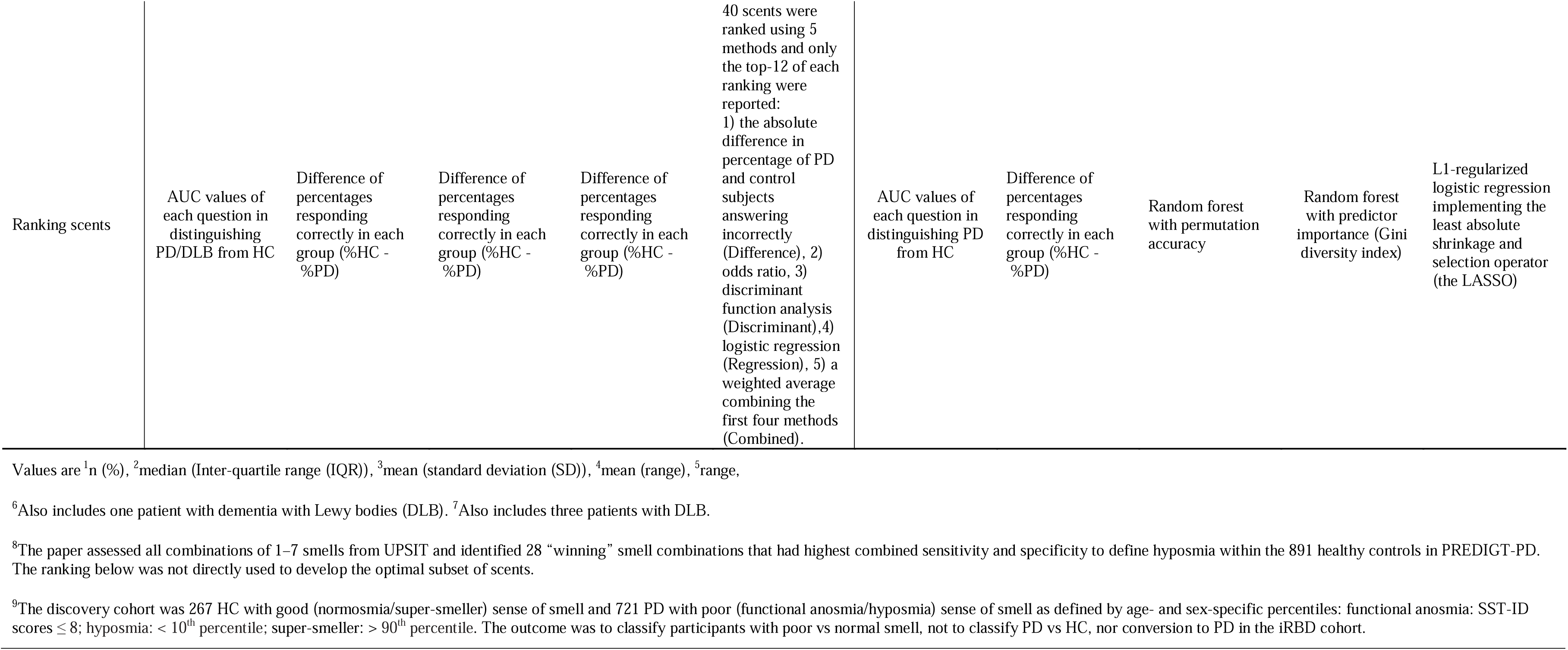
Comparison of cohorts and methods of this study and eight published studies.

**Supplemental Table 3:** Relationship between the whole SST-ID score (DeNoPa) and the whole UPSIT score (Ottawa Trial and PROBE) with age, sex, and diagnostic groups.

|  | DeNoPa |  |  | Ottawa Trial |  |  | PROBE |  |  |
| --- | --- | --- | --- | --- | --- | --- | --- | --- | --- |
| Variable | Beta | 95% CI<sup>1</sup> | p-value | Beta | 95% CI<sup>1</sup> | p-value | Beta | 95% CI<sup>1</sup> | p-value |
| Age | -0.05 | -0.09, -0.01 | **0.022** | -0.22 | -0.31, -0.14 | **<0.001** | -0.21 | -0.31, -0.12 | **<0.001** |
| Sex |  |  |  |  |  |  |  |  |  |
| Female | — | — |  | — | — |  | — | — |  |
| Male | -0.66 | -1.4, 0.06 | 0.073 | -2.9 | -4.6, -1.2 | **<0.001** | -1.9 | -3.9, 0.07 | 0.06 |
| Group |  |  |  |  |  |  |  |  |  |
| HC | — | — |  | — | — |  | — | — |  |
| PD/DLB | -5 | -5.7, -4.3 | **<0.001** | -13 | -14, -11 | **<0.001** | -13 | -15, -11 | **<0.001** |
| MSA/PSP | -1.4 | -3.3, 0.51 | 0.2 | -1.8 | -6.5, 2.9 | 0.5 | -6.4 | -9.2, -3.7 | **<0.001** |

<sup>1</sup> CI = Confidence Interval

## REFERENCES

[1] Haehner A, Hummel T, Reichmann H. Olfactory loss in Parkinson’s disease. Parkinsons Dis. 2011:450939.

[2] Ross GW, Petrovitch H, Abbott RD, et al. Association of olfactory dysfunction with risk for future Parkinson’s disease. Ann Neurol. 2008 Feb;63(2):167–73.

[3] Fereshtehnejad SM, Yao C, Pelletier A, Montplaisir JY, Gagnon JF, Postuma RB. Evolution of prodromal Parkinson’s disease and dementia with Lewy bodies: a prospective study. Brain. 2019 Jul 1;142(7):2051–2067.

[4] McKinnon JH, Demaerschalk BM, Caviness JN, Wellik KE, Adler CH, Wingerchuk DM. Sniffing out Parkinson disease: can olfactory testing differentiate parkinsonian disorders? Neurologist. 2007 Nov;13(6):382–5.

[5] Nalls MA, McLean CY, Rick J, et al. Diagnosis of Parkinson’s disease on the basis of clinical and genetic classification: a population-based modelling study. Lancet Neurol. 2015 Oct;14(10):1002–9.

[6] Bestwick JP, Auger SD, Simonet C, et al. Improving estimation of Parkinson’s disease risk-the enhanced PREDICT-PD algorithm. NPJ Parkinsons Dis. 2021 Apr 1;7(1):33.

[7] Heinzel S, Berg D, Gasser T, Chen H, Yao C, Postuma RB; MDS Task Force on the Definition of Parkinson’s Disease. Update of the MDS research criteria for prodromal Parkinson’s disease. Mov Disord. 2019 Oct;34(10):1464–1470.

[8] Schlossmacher MG, Tomlinson JJ, Santos G, et al. Modelling idiopathic Parkinson disease as a complex illness can inform incidence rate in healthy adults: the PR EDIGT score. Eur J Neurosci. 2017 Jan;45(1):175–191.

[9] Li J, Mestre TA, Mollenhauer B, et al. Evaluation of the PREDIGT score’s performance in identifying newly diagnosed Parkinson’s patients without motor examination. NPJ Parkinsons Dis. 2022 Jul 29;8(1):94.

[10] Doty RL. Psychophysical testing of smell and taste function, Handbook of Clinical Neurology, 164 (2019), pp. 229–246.

[11] Hummel T, Sekinger B, Wolf SR, Pauli E, Kobal G. ’Sniffin’ sticks’: olfactory performance assessed by the combined testing of odor identification, odor discrimination and olfactory threshold. Chem Senses. 1997 Feb;22(1):39–52.

[12] Rumeau C, Nguyen DT, Jankowski R. How to assess olfactory performance with the Sniffin’ Sticks test(®). Eur Ann Otorhinolaryngol Head Neck Dis. 2016 Jun;133(3):203–6.

[13] Boesveldt S, Verbaan D, Knol DL, et al. (2008) A comparative study of odor identification and odor discrimination deficits in Parkinson’s disease. Mov Disord 23: 1984– 1990.

[14] Hawkes CH, Shephard BC, Daniel SE. Olfactory dysfunction in Parkinson’s disease. J Neurol Neurosurg Psychiatry. 1997 May;62(5):436–46.

[15] Bohnen NI, Gedela S, Kuwabara H, et al. Selective hyposmia and nigrostriatal dopaminergic denervation in Parkinson’s disease. J Neurol. 2007 Jan;254(1):84–90.

[16] Morley JF, Cohen A, Silveira-Moriyama L, et al. Optimizing olfactory testing for the diagnosis of Parkinson’s disease: item analysis of the university of Pennsylvania smell identification test. NPJ Parkinsons Dis. 2018 Jan 15;4:2.

[17] Joseph T, Auger SD, Peress L, et al. Screening performance of abbreviated versions of the UPSIT smell test. J Neurol. 2019 Aug;266(8):1897–1906.

[18] Casjens S, Eckert A, Woitalla D, et al. Diagnostic value of the impairment of olfaction in Parkinson’s disease. PLoS One 2013;8:e64735.

[19] Mahlknecht P, Pechlaner R, Boesveldt S, et al. Optimizing odor identification testing as quick and accurate diagnostic tool for Parkinson’s disease. Mov Disord 2016;31:1408–1413.

[20] Lo C, Arora S, Ben-Shlomo Y, et al. Olfactory Testing in Parkinson Disease and REM Behavior Disorder: A Machine Learning Approach. Neurology. 2021 Apr 13;96(15):e2016–e2027.

[21] Double KL, Rowe DB, Hayes M, et al. Identifying the pattern of olfactory deficits in Parkinson disease using the brief smell identification test. Arch Neurol. 2003 Apr;60(4):545–9.

[22] Chou KL, Bohnen NI. Performance on an Alzheimer-selective odor identification test in patients with Parkinson’s disease and its relationship with cerebral dopamine transporter activity. Parkinsonism Relat Disord. 2009 Nov;15(9):640–3.

[23] Gerkin RC, Adler CH, Hentz JG, et al. Improved diagnosis of Parkinson’s disease from a detailed olfactory phenotype. Ann Clin Transl Neurol. 2017 Sep 8;4(10):714–721.

[24] Auger SD, Kanavou S, Lawton M, et al. Testing Shortened Versions of Smell Tests to Screen for Hyposmia in Parkinson’s Disease. Mov Disord Clin Pract. 2020 Mar 21;7(4):394–398.

[25] Vaswani PA, Morley JF, Jennings D, Siderowf A, Marek K; PARS Investigators. Predictive value of abbreviated olfactory tests in prodromal Parkinson disease. NPJ Parkinsons Dis. 2023 Jun 29;9(1):103.

[26] Cohen JF, Korevaar DA, Altman DG, et al. STARD 2015 guidelines for reporting diagnostic accuracy studies: explanation and elaboration. BMJ Open. 2016 Nov 14;6(11): e012799.

[27] Mollenhauer B, Trautmann E, Sixel-Döring F, et al. Nonmotor and diagnostic findings in subjects with de novo Parkinson disease of the DeNoPa cohort. Neurology. 2013 Oct 1;81(14):1226–34.

[28] Diagnostic and Prognostic Biomarkers in Parkinson Disease. https://www.ninds.nih.gov/health-information/clinical-trials/diagnostic-and-prognostic-biomarkers-parkinson-disease

[29] Hughes AJ, Daniel SE, Kilford L, Lees AJ. Accuracy of clinical diagnosis of idiopathic Parkinson’s disease. A clinico-pathological study of 100 cases. J. Neurol. Neurosurg. Psychiatry 55, 181–184 (1992).

[30] Postuma RB, Berg D, Stern M, Poewe W, Olanow CW, Oertel W, et al. MDS clinical diagnostic criteria for Parkinson’s disease. Mov Disord. 2015 Oct;30(12):1591–601.

[31] Cumming G. (2011). Understanding The New Statistics: Effect Sizes, Confidence Intervals, and Meta-Analysis (1st ed.). Routledge. 10.4324/9780203807002

[32] Hummel T, Kobal G, Gudziol H, Mackay-Sim A. Normative data for the "Sniffin’ Sticks" including tests of odor identification, odor discrimination, and olfactory thresholds: an upgrade based on a group of more than 3,000 subjects. Eur Arch Otorhinolaryngol. 2007 Mar;264(3):237–43.

[33] Robin X, Turck N, Hainard A, et al (2011). “pROC: an open-source package for R and S+ to analyze and compare ROC curves.” BMC Bioinformatics, 12, 77. (version 1.18.5)

[34] Youden WJ. Index for rating diagnostic tests. Cancer. 1950 Jan;3(1):32–5.

[35] Ramsay JO, Wiberg M, Li J. (2020). Full Information Optimal Scoring. Journal of Educational and Behavioral Statistics, 45(3), 297–315.

[36] [36] Ho JW, Tumkaya T, (2020). dabestr: Data Analysis using Bootstrap-Coupled Estimation. https://cran.r-project.org/web/packages/dabestr/index.html (version 0.3.0)

[37] Ramsay JO, Li J, Wiberg M, Wallmark J, Graves S. TestGardener: Optimal Analysis of Test and Rating Scale Data. https://cran.r-project.org/web/packages/TestGardener/index.html (version 3.2.6)

[38] Wickham H (2016). ggplot2: Elegant Graphics for Data Analysis. Springer-Verlag New York. ISBN 978-3-319-24277-4, https://ggplot2.tidyverse.org/. (version 3.4.4)

[39] Ramsay JO, Hooker G, Graves S. fda: Functional Data Analysis. https://cran.r-project.org/web/packages/fda/index.html (version 6.1.4)

[40] Lang AE, Siderowf AD, Macklin EA, et al. Trial of Cinpanemab in Early Parkinson’s Disease. N Engl J Med. 2022 Aug 4;387(5):408–420.

[41] Pagano G, Taylor KI, Anzures-Cabrera J, et al. Trial of Prasinezumab in Early-Stage Parkinson’s Disease. N Engl J Med. 2022 Aug 4;387(5):421–432.

[42] Jensen PH, Schlossmacher MG, Stefanis L. Who Ever Said It Would Be Easy? Reflecting on Two Clinical Trials Targeting α-Synuclein. Mov Disord. 2023 Mar;38(3):378–384.

[43] Jennings D, Siderowf A, Stern M, et al. (2014) Imaging prodromal Parkinson disease: the Parkinson Associated Risk Syndrome study. Neurology 83:1739–1746.

[44] The Lancet. What next in Parkinson’s disease? Lancet. 2024 Jan 20;403(10423):219.

[45] [45] Mollenhauer B, Li J, Schlossmacher MG (2023). Persistent Hyposmia as Surrogate for α-Synuclein-Linked Brain Pathology. medRxiv https://www.medrxiv.org/content/10.1101/2023.12.19.23300164v2

[46] Stefani A, Iranzo A, Holzknecht E, et al. Alpha-synuclein seeds in olfactory mucosa of patients with isolated REM sleep behaviour disorder. Brain. 2021 May 7;144(4):1118–1126.

[47] Tomlinson JJ, Shutinoski B, Dong L, et al. Holocranohistochemistry enables the visualization of α-synuclein expression in the murine olfactory system and discovery of its systemic anti-microbial effects. J Neural Transm (Vienna). 2017 Jun;124(6):721–738.

[48] Martin-Lopez E, Vidyadhara DJ, Liberia T, et al. α-Synuclein Pathology and Reduced Neurogenesis in the Olfactory System Affect Olfaction in a Mouse Model of Parkinson’s Disease. J Neurosci. 2023 Feb 8;43(6):1051–1071.

[49] Chen F, Liu W, Liu P, et al. α-Synuclein aggregation in the olfactory bulb induces olfactory deficits by perturbing granule cells and granular-mitral synaptic transmission. NPJ Parkinsons Dis. 2021 Dec 13;7(1):114.

[50] Petit GH, Berkovich E, Hickery M, et al. Rasagiline ameliorates olfactory deficits in an alpha-synuclein mouse model of Parkinson’s disease. PLoS One. 2013;8(4):e60691.

[51] Fleming SM, Tetreault NA, Mulligan CK, Hutson CB, Masliah E, Chesselet MF. Olfactory deficits in mice overexpressing human wildtype alpha-synuclein. Eur J Neurosci. 2008 Jul;28(2):247–56.

