## Supplemental Figures for "Development of a Simplified Smell Test to Identify Patients with Typical Parkinson’s as Informed by Multiple Cohorts, Machine Learning and External Validation"

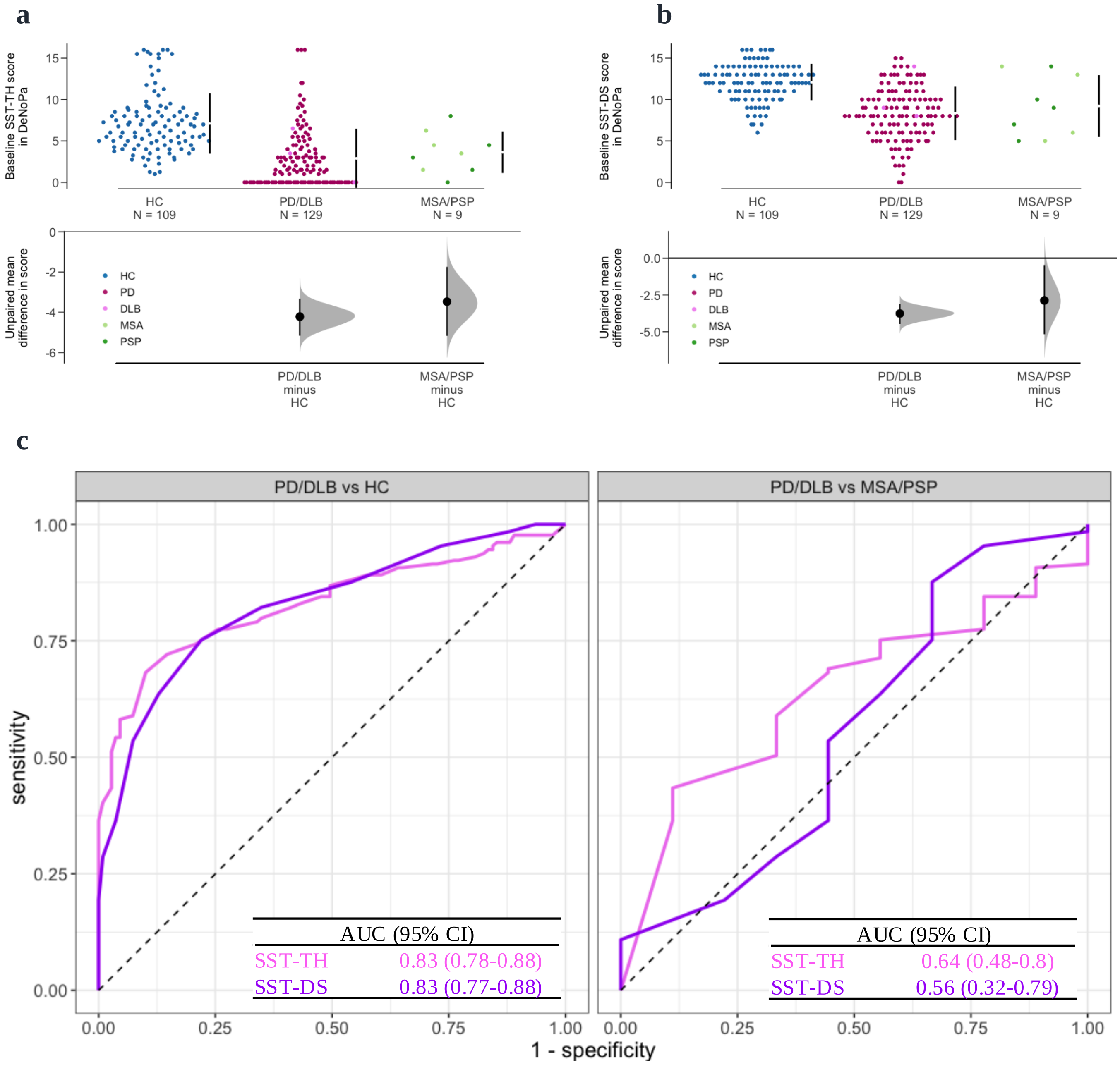


**Supplemental Figure 1: Distribution of Sniffin' Sticks Threshold (SST-TH) and Discrimination (SST-DS) scores for each subject group in the DeNoPa Study at baseline and their ROC curves for group classification.**

The Cummings estimation plots (a, b) were used to illustrate and compare smell test score distributions in each group: (a) SST-TH, (b) SST-DS. Each data point in the upper panels represents the score of one participant, and colors represent different groups and diagnosis as shown in the legend. The vertical lines in the upper panels represent the conventional mean ± standard deviation error bars. The lower panels show the mean group difference (the effect size) and its 95% confidence interval (CI) estimated by bias-corrected and accelerated bootstrap, using HC as the reference group. Panel (c) shows the ROC curves of each smell test (indicated by color) to distinguish PD/DLB versus HC (left) and PD versus OND (right). HC = healthy control. PD = Parkinson disease. DLB = dementia with Lewy bodies. MSA = multiple system atrophy. PSP = progressive supranuclear palsy. ROC = receiver operating characteristic. AUC = area under the ROC curve.

a

b

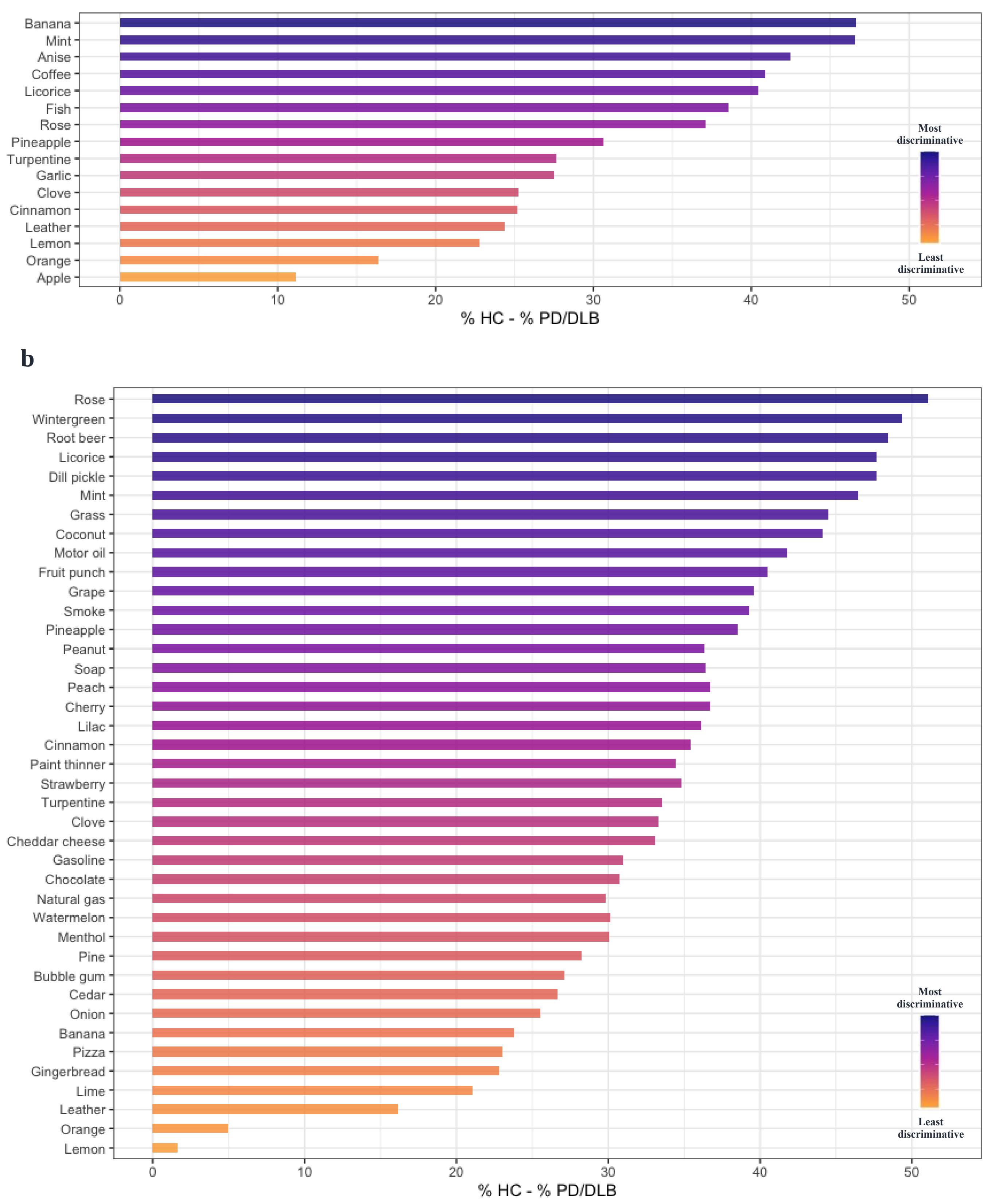


**Supplemental Figure 2: Percentage differences of correct scent identification between HC and PD/DLB groups (% HC - % PD/DLB) in the DeNoPa (a) and Ottawa Trial (b) cohorts.**
The scents are ordered in descending orders of their mean single-scent AUC value (see Figure 3 (a) and (c)); the color of each scent changes gradually from the most discriminative to the least discriminative odorant, as indicated by the legend.

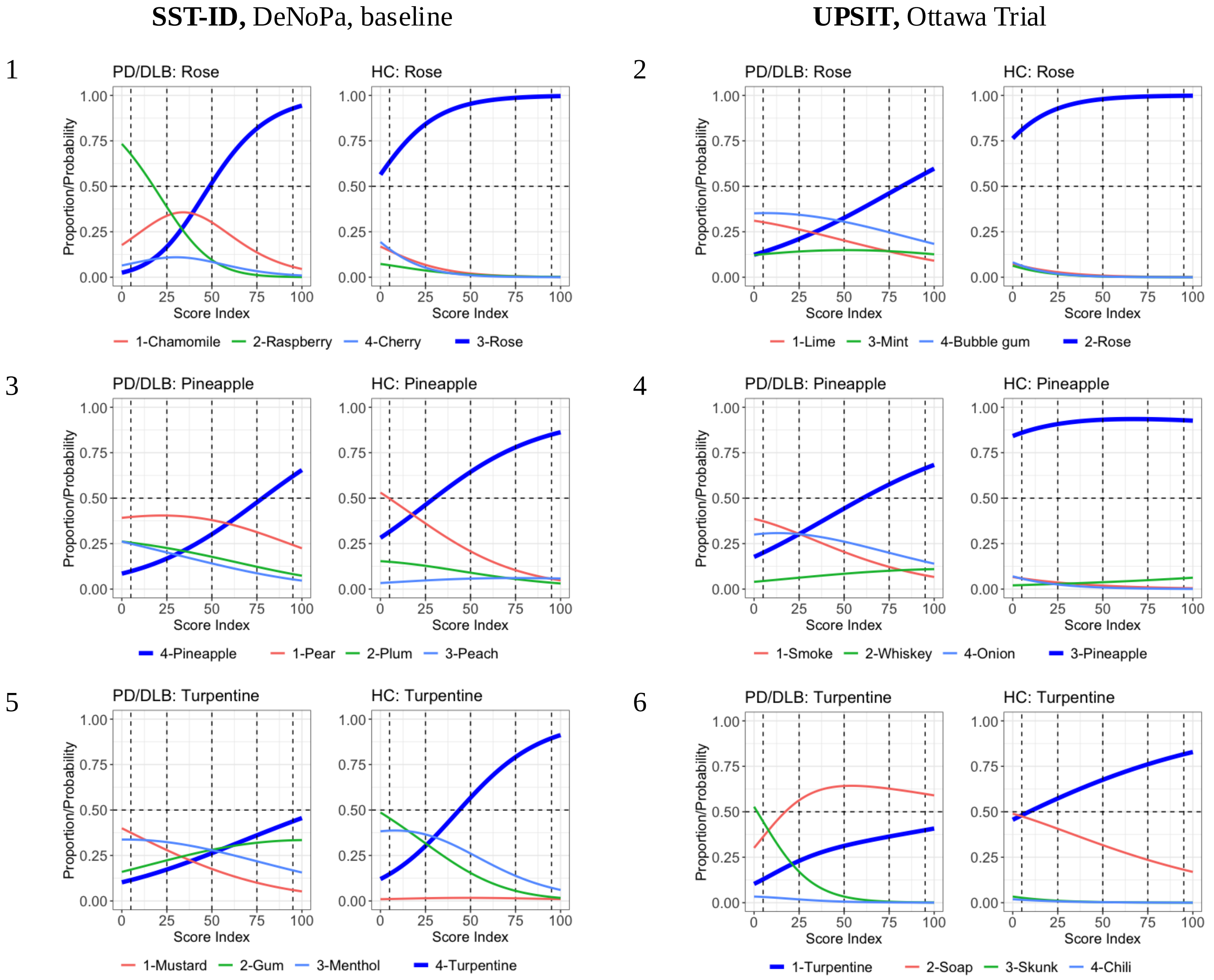


**Supplemental Figure 3: Influence of distractors in the multiple-choice smell tests: the remaining 6 scents shared by UPSIT and SST-ID (1/2).**

Panels with odd numbers show the Item Characteristic Curves (ICCs) of six SST-ID scents, and panels with even numbers show the ICCs of the corresponding UPSIT scents. In each figure, the left panels are the ICCs using data of the PD/DLB patients, and the right panels are corresponding to healthy controls. The x-axis is transformed score indices (percentage rank of the SST-ID score) within the corresponding group. The y-axis is the probability of choosing each option at a particular score index. The correct option of each item is highlighted using the thicker blue curves. Numbers in the color legends are the option indices. The horizontal dashed lines represent 50% probability. The vertical dashed lines represent five quantiles (5%, 25%, 50%, 75%, and 95%).

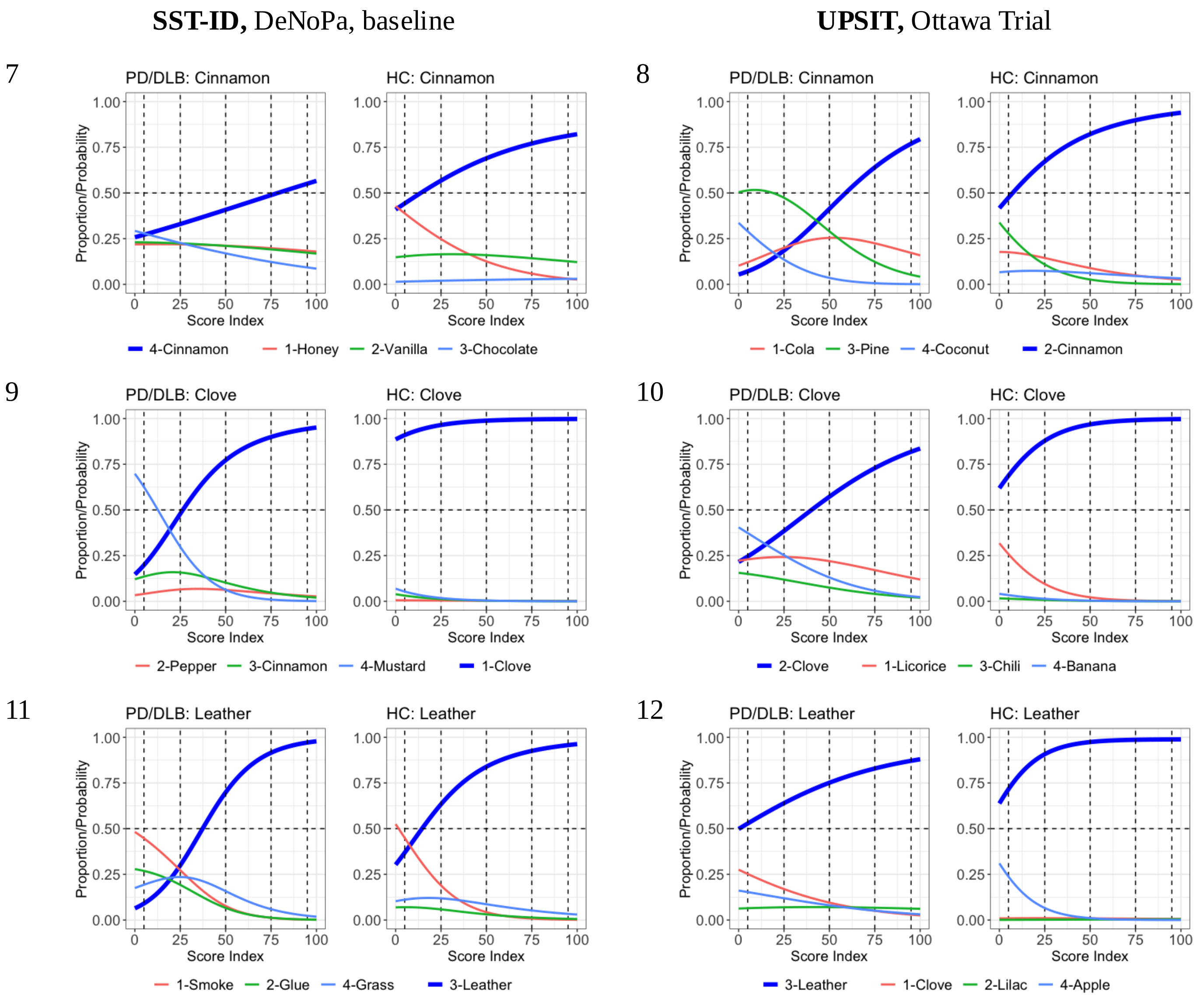


**Supplemental Figure 3: Influence of distractors in the multiple-choice smell tests: the remaining 6 scents shared by USPIT and SST-ID (2/2).**

Panels with odd numbers show the Item Characteristic Curves (ICCs) of six SST-ID scents, and panels with even numbers show the ICCs of the corresponding UPSIT scents. In each figure, the left panels are the ICCs using data of the PD/DLB patients, and the right panels are corresponding to healthy controls. The x-axis is transformed score indices (percentage rank of the SST-ID score) within the corresponding group. The y-axis is the probability of choosing each option at a particular score index. The correct option of each item is highlighted using the thicker blue curves. Numbers in the color legends are the option indices. The horizontal dashed lines represent 50% probability. The vertical dashed lines represent five quantiles (5%, 25%, 50%, 75%, and 95%).

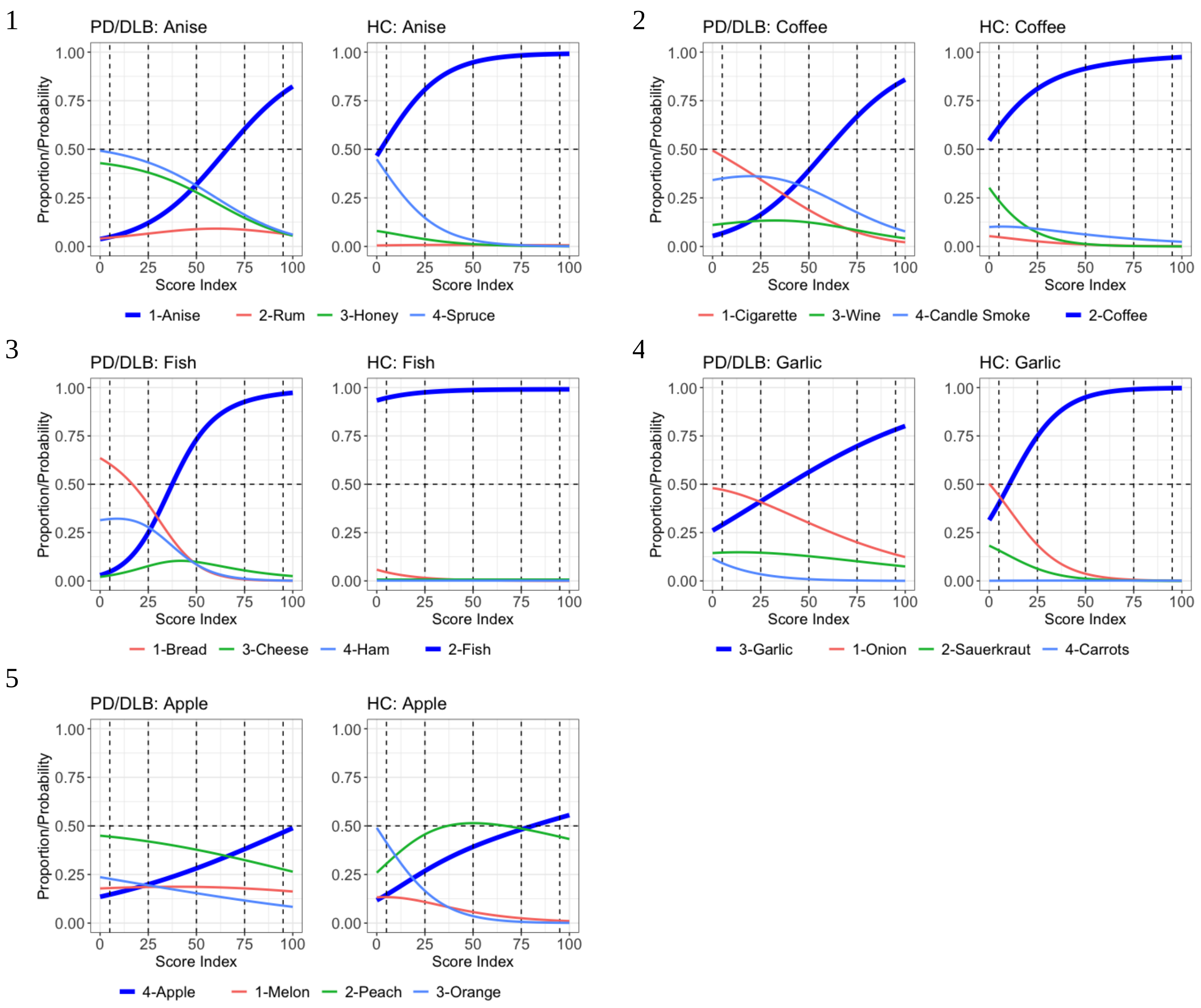


**Supplemental Figure 4: Item Characteristic Curves (ICCs) of the remaining SST-ID scents using baseline data from the DeNoPa Cohort.**

In each panel, the left panels are the ICCs using data of the PD/DLB patients, and the right panels are corresponding to healthy controls. The x-axis is transformed score indices (percentage rank of the SST-ID score) within PD and HC group, respectively. The y-axis is the probability of choosing each option at a particular score index. The correct option of each item is highlighted using the thicker blue curves. Numbers in the color legends are the option indices. The horizontal dashed lines represent 50% probability. The vertical dashed lines represent five quantiles (5%, 25%, 50%, 75%, and 95%).

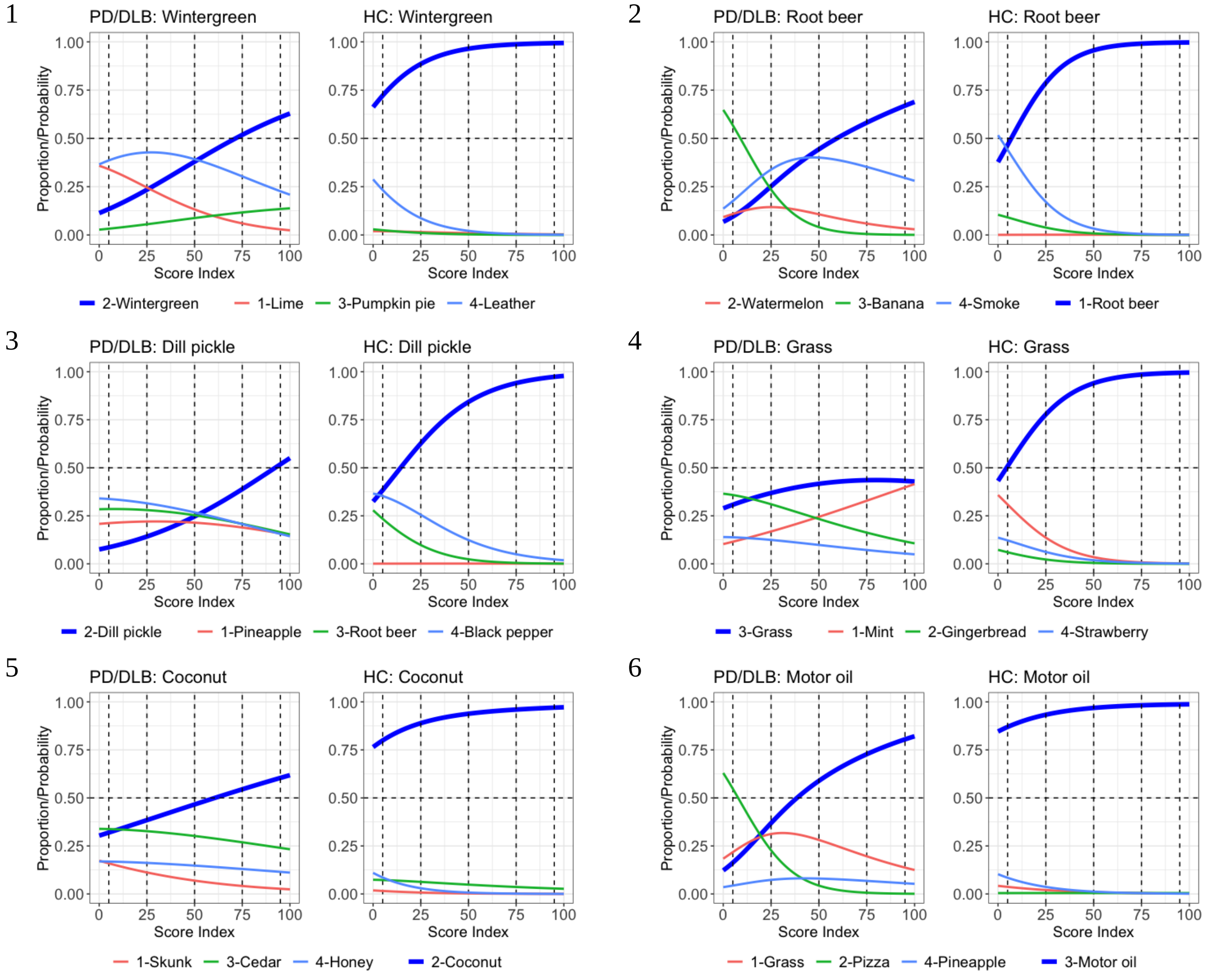


**Supplemental Figure 5: Item Characteristic Curves (ICCs) of the remaining UPSIT scents using the Ottawa Trial cohort (1/5).**

In each panel, the left panels are the ICCs using data of the PD/DLB patients, and the right panels are corresponding to healthy controls. The x-axis is transformed score indices (percentage rank of the SST-ID score) within PD and HC group, respectively. The y-axis is the probability of choosing each option at a particular score index. The correct option of each item is highlighted using the thicker blue curves. Numbers in the color legends are the option indices. The horizontal dashed lines represent 50% probability. The vertical dashed lines represent five quantiles (5%, 25%, 50%, 75%, and 95%).

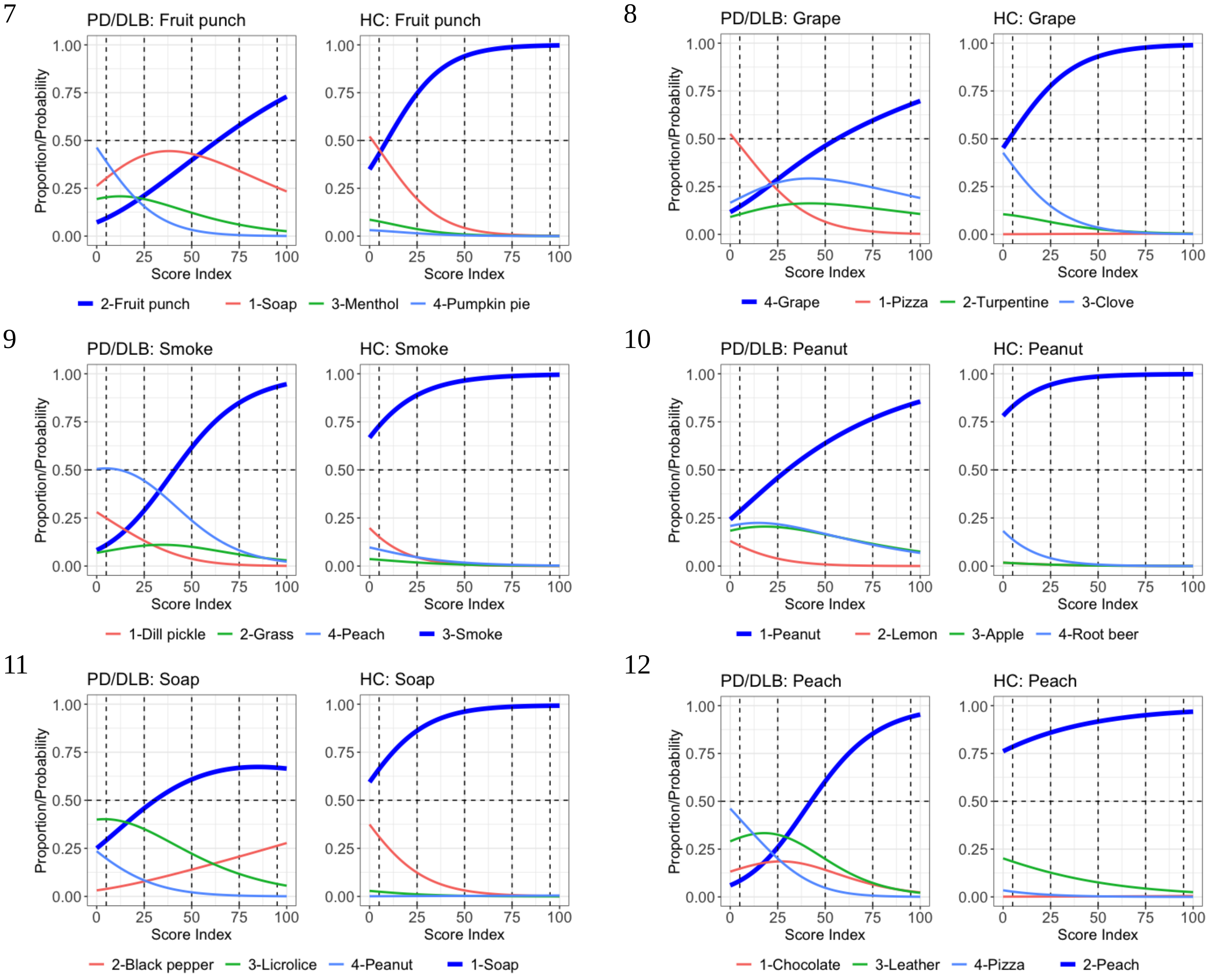


**Supplemental Figure 5: Item Characteristic Curves (ICCs) of the remaining UPSIT scents using the Ottawa Trial cohort (2/5).**

In each panel, the left panels are the ICCs using data of the PD/DLB patients, and the right panels are corresponding to healthy controls. The x-axis is transformed score indices (percentage rank of the SST-ID score) within PD and HC group, respectively. The y-axis is the probability of choosing each option at a particular score index. The correct option of each item is highlighted using the thicker blue curves. Numbers in the color legends are the option indices. The horizontal dashed lines represent 50% probability. The vertical dashed lines represent five quantiles (5%, 25%, 50%, 75%, and 95%).

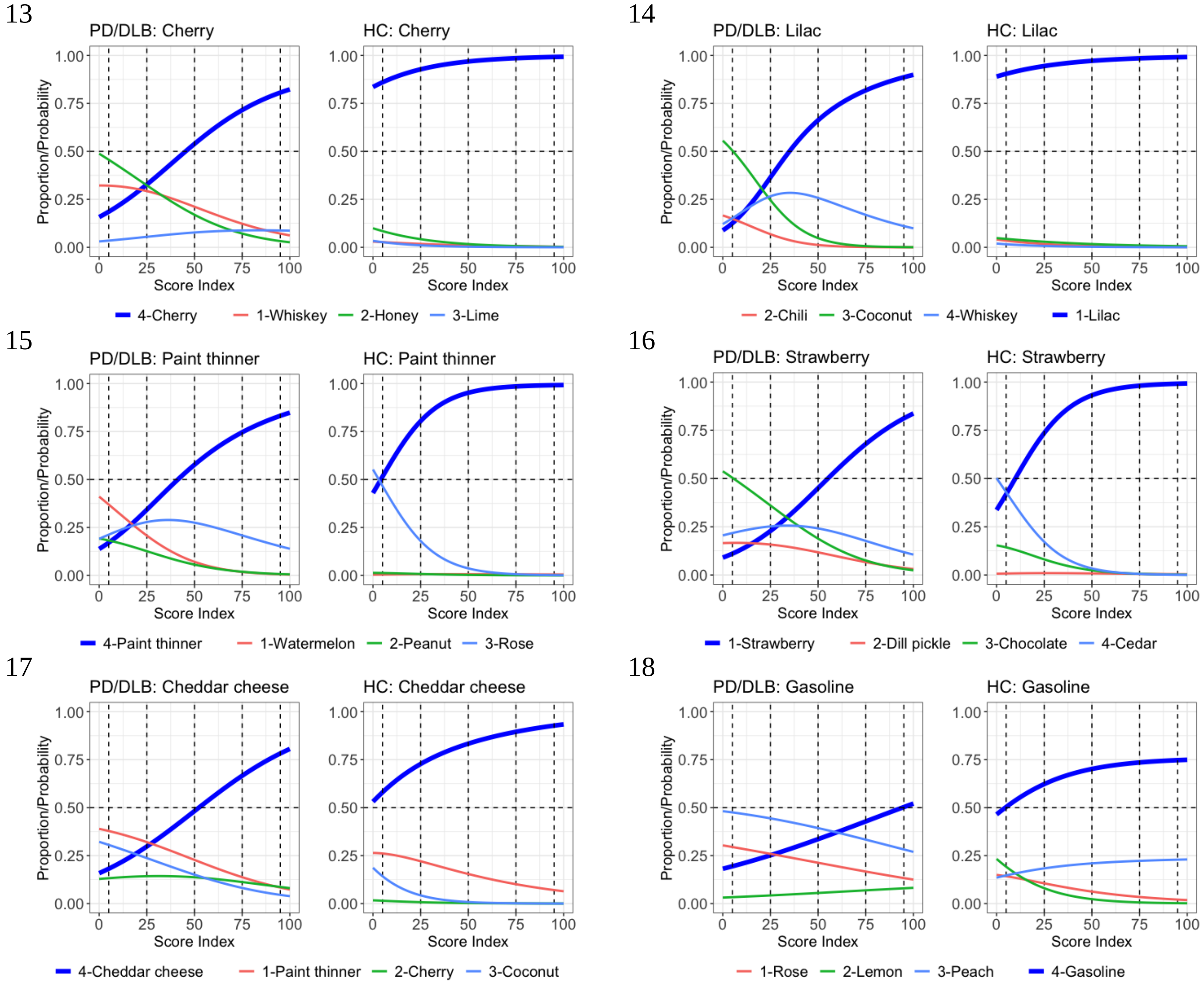


**Supplemental Figure 5: Item Characteristic Curves (ICCs) of the remaining UPSIT scents using the Ottawa Trial cohort (3/5).**

In each panel, the left panels are the ICCs using data of the PD/DLB patients, and the right panels are corresponding to healthy controls. The x-axis is transformed score indices (percentage rank of the SST-ID score) within PD and HC group, respectively. The y-axis is the probability of choosing each option at a particular score index. The correct option of each item is highlighted using the thicker blue curves. Numbers in the color legends are the option indices. The horizontal dashed lines represent 50% probability. The vertical dashed lines represent five quantiles (5%, 25%, 50%, 75%, and 95%).

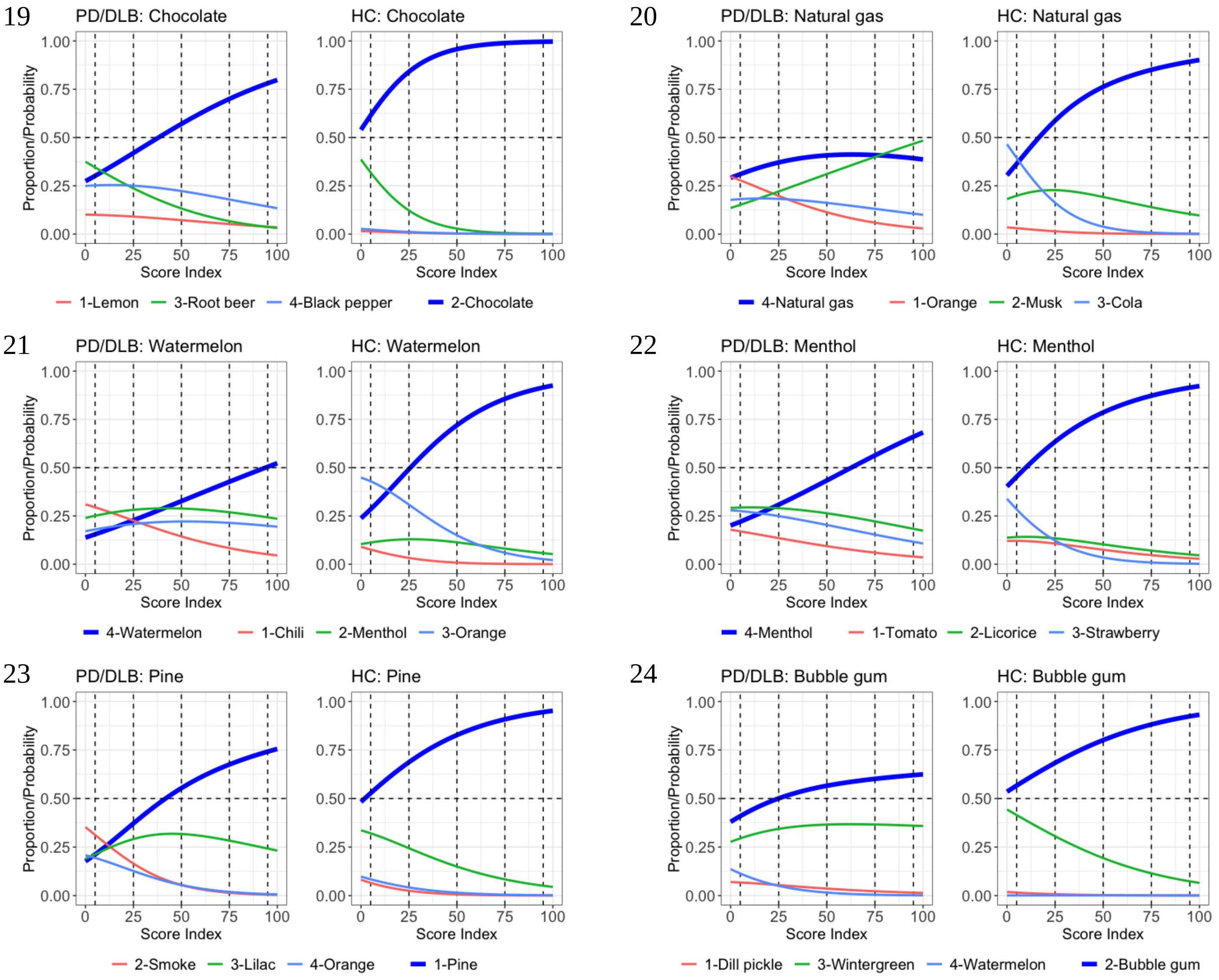


**Supplemental Figure 5: Item Characteristic Curves (ICCs) of the remaining UPSIT scents using the Ottawa Trial cohort (4/5).**

In each panel, the left panels are the ICCs using data of the PD/DLB patients, and the right panels are corresponding to healthy controls. The x-axis is transformed score indices (percentage rank of the SST-ID score) within PD and HC group, respectively. The y-axis is the probability of choosing each option at a particular score index. The correct option of each item is highlighted using the thicker blue curves. Numbers in the color legends are the option indices. The horizontal dashed lines represent 50% probability. The vertical dashed lines represent five quantiles (5%, 25%, 50%, 75%, and 95%).

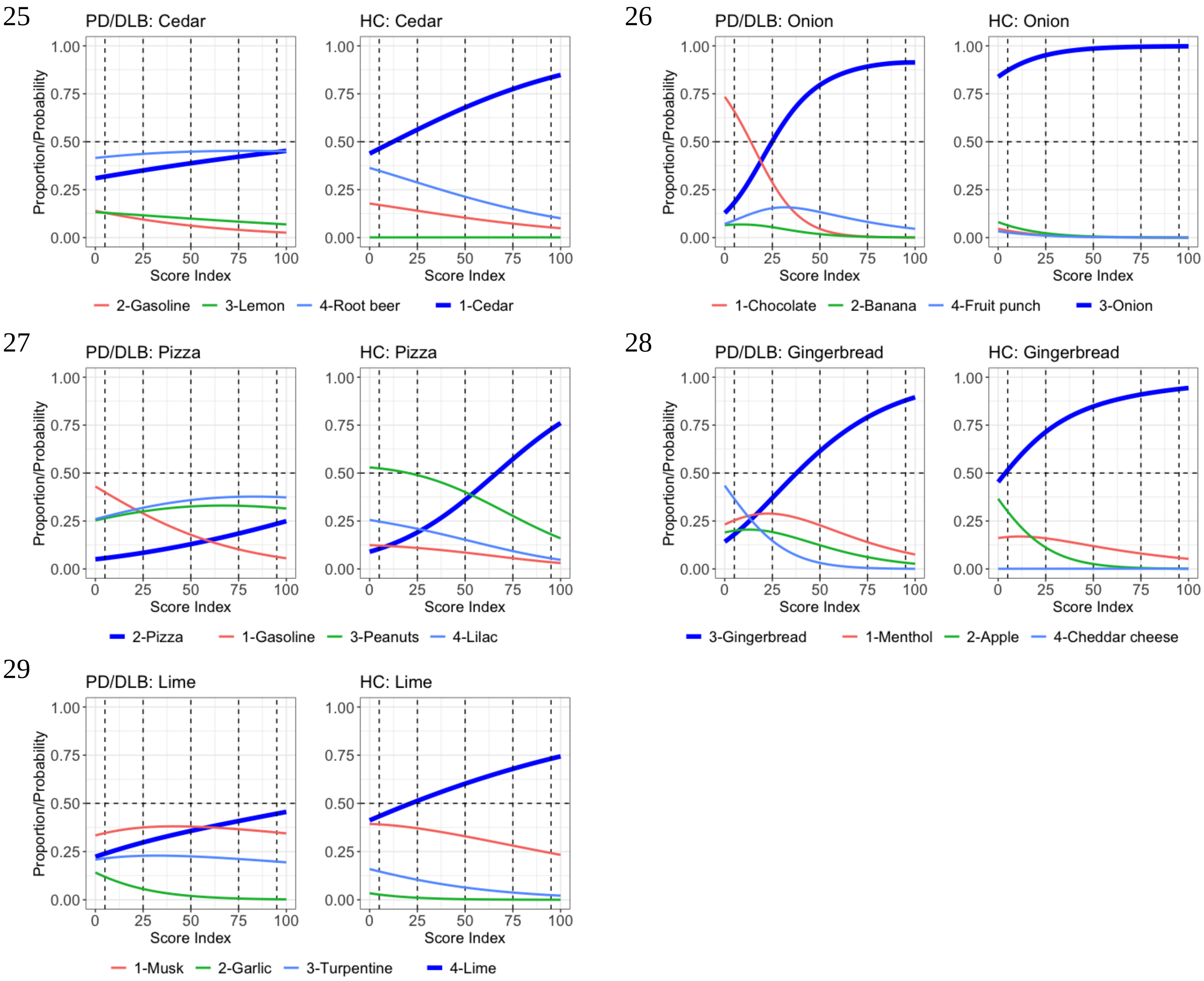


**Supplemental Figure 5: Item Characteristic Curves (ICCs) of the remaining UPSIT scents using the Ottawa Trial cohort (5/5).**

In each panel, the left panels are the ICCs using data of the PD/DLB patients, and the right panels are corresponding to healthy controls. The x-axis is transformed score indices (percentage rank of the SST-ID score) within PD and HC group, respectively. The y-axis is the probability of choosing each option at a particular score index. The correct option of each item is highlighted using the thicker blue curves. Numbers in the color legends are the option indices. The horizontal dashed lines represent 50% probability. The vertical dashed lines represent five quantiles (5%, 25%, 50%, 75%, and 95%).

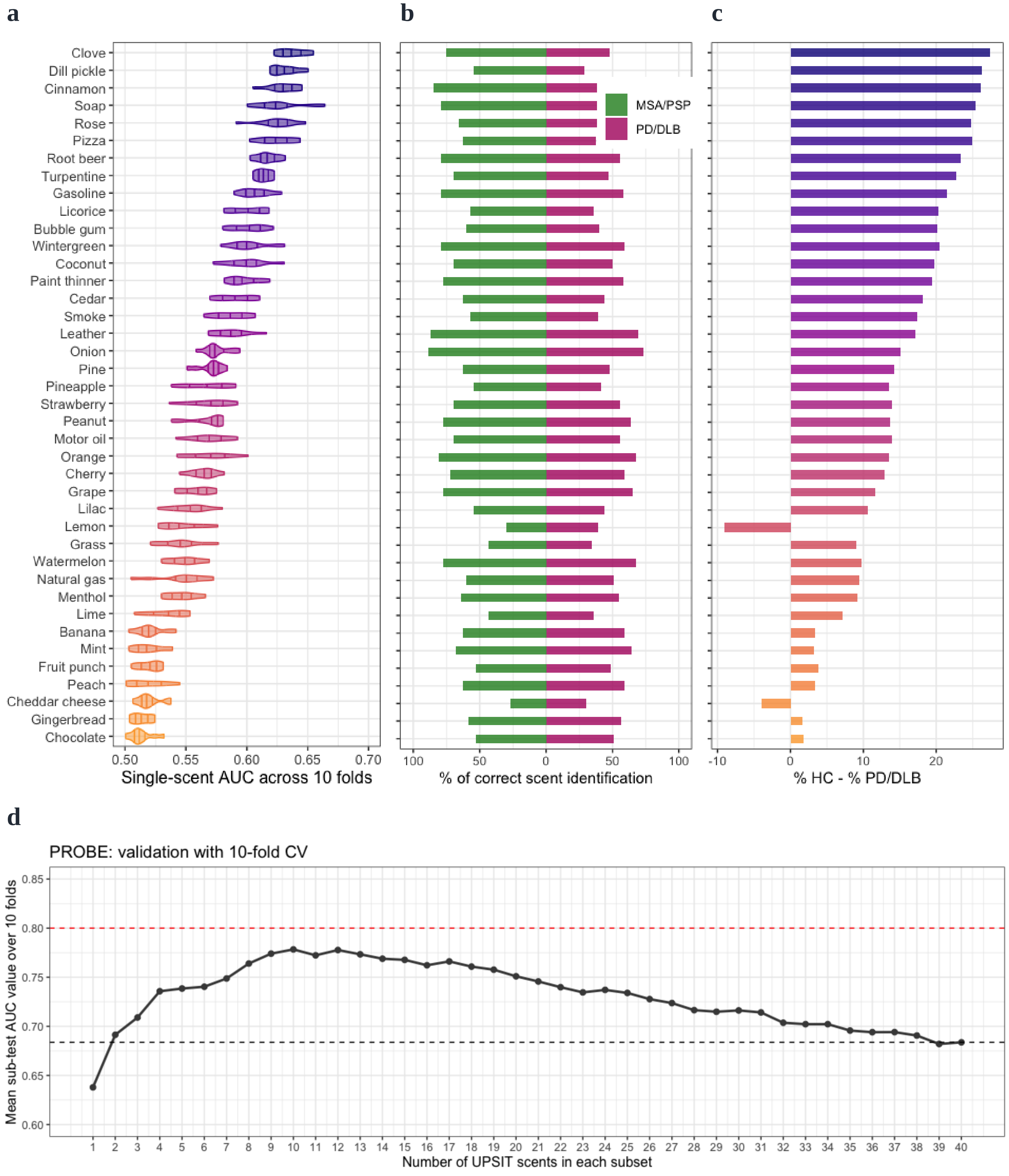


**Supplemental Figure 6: Range of performances for UPSIT scents in differentiating PD/DLB from MSA/PSP subjects in the PROBE cohort and AUC values for numerical subsets of odorants in group classification.**

Panel (a) illustrates distribution of AUC values of each scent across 10-fold cross-validation using violin plots, with 25%, 50%, and 75% quantile lines. The scents are ordered in descending order of their mean single-scent AUC value. The color of each scent changes gradually from the most to the least discriminative odorant, as indicated by the legend. Panel (b) shows the percentage of subjects correctly identifying each scent within the MSA/PSP and PD/DLB groups, and panel (c) shows the percentage differences between the two groups. Scents in panels (b) and (c) follow the same rank order as in panel (a). In panel (d), the x-axis is the number of scents included for each subset, with individual points representing average AUC values for the validation set across ten CV folds gathered in PROBE. The black horizontal, dashed line indicates the corresponding AUC values of the whole test (= 40 scents). The red horizontal, dashed line indicates AUC = 0.8 as a predetermined reference line.
